# Comprehensive real-world utilization, treatment trajectories, and outcomes of immune checkpoint inhibitors across cancer types: the multicenter FAIR-ICI cohort

**DOI:** 10.64898/2026.11.02.26367076

**Authors:** Linh T. Ha, Fabienne Ver Donck, Iege Bassez, Clara L. Oeste, Christof Vulsteke, Danielle Delombaerde, Dries Hens, Hans Prenen, Lieselot Croes, Astrid De Meulenaere, Philip R. Debruyne, Marthe Verhaert, Sandrine Aspeslagh, Nele Brusselaers, Annelies Verbiest

**Author notes:** **Corresponding author:** Annelies Verbiest, MD, PhD, Department of Medical Oncology, Multidisciplinary Oncology Center Antwerp, Antwerp University Hospital, Edegem, Belgium.

## Abstract

**Background:** Immune checkpoint inhibitors (ICIs) have transformed cancer treatment, yet evidence on their utilization, treatment trajectories, and effectiveness in routine clinical practice remains limited. We characterized real-world ICI use and outcomes across cancer types using the multicenter FAIR-ICI cohort and explored differences between real-world and pivotal trial outcomes.

**Methods:** We conducted a retrospective cohort study of adults receiving ICIs across four Belgian hospitals. Structured and unstructured electronic health record data were harmonized using the Observational Medical Outcomes Partnership Common Data Model (OMOP-CDM) and natural language processing. We evaluated overall survival (OS), time on treatment (ToT), time to next treatment (TTNT), ICI discontinuation (>12-week treatment gap without re-initiation), and post-discontinuation survival. OS was descriptively compared with pivotal trials, and prognostic factors were assessed using multivariable Cox regression. An interactive public dashboard was developed to enable exploration of cohort characteristics, treatment patterns, and outcomes.

**Results:** Among 3,231 ICI-treated patients, 2,514 (77.8%) had metastatic disease. Among metastatic patients, 568 (22.6%) discontinued ICI without subsequent systemic therapy. Median post-discontinuation OS was 21.4 months (95% CI 16.5–37.2), with 49.1% alive at 24 months. ToT and TTNT were generally shorter than OS, particularly in melanoma (median OS 78 months; ToT 7 months; TTNT 26 months). Real-world survival was frequently less favorable than in pivotal trials. ECOG ≥2 was associated with poorer OS than ECOG 0–1 (adjusted HR 2.18, 95% CI 1.82–2.60). In NSCLC patients receiving pembrolizumab plus chemotherapy, survival with ECOG 0–1 more closely approximated trial outcomes, whereas survival with ECOG ≥2 was substantially poorer.

**Conclusions:** Real-world ICI treatment trajectories extend beyond active treatment, with some patients experiencing prolonged survival after discontinuation. Differences between real-world and trial outcomes highlight the importance of patient characteristics, particularly performance status, when interpreting trial evidence in routine care. FAIR-ICI provides a scalable framework for evaluating real-world immunotherapy effectiveness, treatment trajectories, and future investigations of treatment-related toxicity.

**Highlights:**

- FAIR-ICI studied 3,231 patients across four Belgian hospitals and provides an interactive dashboard for subgroup exploration.
- Among metastatic patients, 22.6% stopped ICIs without subsequent systemic therapy, with 49.1% alive at 24 months after stopping.
- Time on treatment and time to next treatment were generally shorter than overall survival.
- Real-world survival was frequently less favorable than in pivotal clinical trials.
- Poor ECOG performance status was strongly associated with shorter survival and may contribute to trial–real-world differences.

## Introduction

Immune checkpoint inhibitors (ICIs) have transformed cancer treatment, improving survival and becoming standard-of-care across multiple malignancies. Since the European Medicines Agency (EMA) approved ipilimumab for metastatic melanoma in 2011^1^, ICI indications have expanded rapidly across tumor types, disease stages, and treatment combinations^2,3^. Recent estimates indicate that approximately 57% of patients with advanced or metastatic cancer may be eligible for ICI treatment ^4^, highlighting the growing role of ICIs in contemporary oncology care.

Belgium, with around 77,000 new cancer diagnoses annually^5^, provides an informative setting for evaluating real-world ICI use. Since 2017, an accelerated reimbursement pathway has enabled access to ICIs immediately following EMA approval, rather than through the conventional Belgian reimbursement procedure^6^, with ICIs currently reimbursed across 23 cancer types^7^. A recent European Comparator Report ranked Belgium among countries with the highest ICI uptake (roughly six standard weekly doses per incident cancer case)^8^. At current Belgian list prices, this corresponds to an estimated drug cost of approximately €10,800 per incident cancer case. As ICI use continues to expand globally, evidence from this setting can inform understanding of real-world treatment patterns, effectiveness, and resource implications across healthcare systems.

Despite widespread use, evidence supporting ICIs remains largely derived from randomized controlled trials (RCTs), whose populations and treatment pathways may not reflect routine clinical practice^9^. Previous studies have estimated that approximately 70% of NSCLC patients were not eligible for pivotal phase III ICI trials^10^. Such differences may contribute to the efficacy–effectiveness gap between clinical trials and real-world populations. Furthermore, RCTs provide limited insight into treatment trajectories, including treatment duration, discontinuation, and transitions to subsequent therapy^11,12^, which are important for understanding clinical decision-making, patient outcomes, and healthcare resource use.

Real-world evidence (RWE) complements clinical trials by evaluating treatment use and outcomes in routine practice^13^. However, comprehensive multicenter studies spanning multiple tumor types remain limited, often constrained by fragmented data sources and insufficient clinical detail^14^. Most studies focus primarily on overall survival (OS), whereas treatment-trajectory endpoints such as time on treatment (ToT) and time to next treatment (TTNT) are less frequently reported. Similarly, little is known about treatment discontinuation patterns, including the proportion of patients discontinuing ICIs without subsequent systemic treatment and their survival.

To address these challenges, the FAIR-ICI cohort was established as a multicenter Belgian real-world dataset integrating structured and unstructured electronic health record (EHR) data within a standardized Observational Medical Outcomes Partnership Common Data Model (OMOP-CDM) framework^15,16^. Natural language processing (NLP) enriches structured data with clinically relevant information extracted from routine records. By combining multicenter patient-level data, standardized data harmonization, and NLP-enriched clinical information, FAIR-ICI enables high-resolution evaluation of ICI utilization, treatment trajectories, and outcomes across routine oncology practice.

In this study, we aimed to (1) assess patient demographics, disease characteristics, and treatment patterns among patients receiving ICIs; (2) estimate real-world outcomes, including OS, TTNT, and ToT; (3) characterize treatment discontinuation patterns and survival following discontinuation; and (4) compare real-world survival with pivotal clinical trials. To facilitate transparent exploration, we additionally developed an interactive dashboard that enables dynamic subgroup analyses and visualization of outcomes. Together, these analyses provide a comprehensive overview of ICI use and effectiveness in routine oncology care and establish FAIR-ICI as a platform for future research on immunotherapy-related toxicity.

## Methods

### Study design and setting

This multicenter, retrospective observational cohort study used routinely collected EHR data from four Belgian hospitals: two academic centers (University Hospital Antwerp and University Hospital Brussels) and two regional hospitals (Maria Middelares Hospital Ghent and AZ Groeninge Hospital Kortrijk). All institutions maintain OMOP-CDM version 5 databases generated through standardized extract–transform–load (ETL) procedures. The study period extended from September 2015 to November 2024 and covered all available data, with start and end dates varying across data partners and data sources (start date ranging from September 2015 to January 2018, and end date from November 2023 to November 2024).

### Data sources and processing

The FAIR-ICI database integrates structured data (hospital cancer registries, medication administration records, laboratory measurements, administrative datasets, and minimal hospital data–mandatory Belgian reporting datasets containing harmonized administrative and diagnostic information) and unstructured clinical text (tumor board reports, consultation notes, pathology reports, radiology reports, and discharge summaries).

Unstructured clinical text was processed using a previously developed NLP pipeline^17^. NLP-derived clinical concepts were mapped to Unified Medical Language System (UMLS) Concept Unique Identifiers (CUIs) and subsequently converted to standardized OMOP concept identifiers using OMOP Athena vocabularies.

Structured and NLP-derived variables were transformed into the OMOP-CDM. The data extraction and processing workflow is illustrated in **Supplementary-Figure S1. Supplementary-Tables S1–S3** provide the datapoint list (including source systems, OMOP tables, and UMLS CUIs), data dictionary (variable definitions, derivation logic, timing, and data sources), and NLP performance metrics, respectively.

Data were pseudonymized before ETL processing.

### Study population

The study population included adult patients who initiated ICI treatment during the study period. The index event was defined as the first ICI administration. Eligible ICIs, available in Belgium during the study period, included PD-1 inhibitors (nivolumab, pembrolizumab, cemiplimab, and dostarlimab), PD-L1 inhibitors (atezolizumab, durvalumab, and avelumab), and CTLA-4 inhibitors (ipilimumab and tremelimumab), either as monotherapy or in approved combination regimens. To make sure we only included new ICI users, patients were required to have at least 183 days of prior observation, except when cancer diagnosis occurred within 21 days before the index.

Patients were followed from the index date until cohort exit, which happened at death, diagnosis of a second primary malignancy, initiation of a new ICI regimen, ICI re-initiation after a gap exceeding 12 weeks, or end of available data.

Cancer type was assigned using structured cancer records and NLP-derived diagnoses, considering reimbursed ICI indications (**Supplementary-Methods S1; Figure S2**). Tumor stage at diagnosis and ICI initiation was determined using TNM staging and metastasis-related information (**Supplementary-Methods S2**).

### Variables and outcomes

Baseline variables included demographic characteristics (age, sex, body mass index [BMI]), lifestyle factors (smoking status and alcohol abuse), Eastern Cooperative Oncology Group (ECOG) performance status, tumor type, and stage at ICI initiation.

Treatment characteristics included ICI initiation year, ICI agent, mechanism of action, and regimen classification. Regimen type was determined according to all systemic anticancer therapies administered on the index date and categorized as ICI monotherapy, dual ICI therapy, ICI plus chemotherapy, ICI plus tyrosine kinase inhibitor (TKI), or other ICI-containing combinations.

Patients were considered to be on treatment until 12 weeks after last ICI administration or initiation of a new systemic antineoplastic regimen, whichever occurred first. A gap exceeding 12 weeks was considered permanent discontinuation of the index treatment episode. Treatment status at data cut off was classified as ongoing treatment, death during treatment, switch to a new systemic anticancer regimen, or permanent discontinuation.

The primary time-to-event outcomes were OS, ToT, and TTNT. OS was defined as the time from the index date to death from any cause, based on linkage to hospital and governmental death records. Patients without a registered death date were censored at the last recorded observation. ToT was defined as the time from the index date to treatment-ending events, including death, initiation of a new systemic anticancer regimen, or permanent discontinuation of the index ICI regimen. TTNT was defined as the time from the index date to initiation of a new systemic treatment regimen not part of the initial combination regimen. The event date was defined as the first administration of the new regimen. Switching to another ICI or addition of chemotherapy, targeted therapy, or hormonal therapy was considered a next treatment. Death was treated as a censoring event.

### Validation

Data quality was assessed through a multilayered validation framework: data intake validation, NLP validation, clinical validation of individual datapoints, and clinical validation of aggregated outputs.

During data intake validation, participating hospitals verified completeness and coverage of source datasets through checks for missing values, date inconsistencies, duplicate records, and compliance with inclusion criteria.

LynxCare Clinical Informatics evaluated NLP-derived variables against human-annotated reference datasets, calculating precision, recall, and F1 scores for each datapoint. Models were iteratively refined until predefined performance targets were achieved (**Supplementary-Table S3**).

Physicians at participating hospitals validated individual datapoints by reviewing concept frequencies, terminology mappings, source attribution, and concordance between structured and unstructured sources. Investigators then reviewed aggregated cohort outputs at site and consortium levels, iteratively assessing summary statistics, phenotype definitions, and variable distributions to identify inconsistencies, refine extraction rules, and update the FAIR-ICI data dictionary. These validation procedures, together with predefined phenotype algorithms, aimed to minimize data extraction errors and misclassification.

The multidisciplinary FAIR-ICI team brought together clinical oncologists, epidemiologists, and clinical informatics specialists from participating hospitals and LynxCare to support data harmonization, validation, analysis, and clinical interpretation.

### Statistical analysis

Baseline characteristics were summarized as medians with interquartile ranges (IQR) for continuous variables and frequencies and percentages for categorical variables. Missing values were retained as a separate category.

OS, ToT, and TTNT were estimated using the Kaplan–Meier method, reporting median times with corresponding 95% confidence intervals (CIs) and survival probabilities at selected time points. Differences between Kaplan– Meier curves were assessed using the log-rank test. To explore factors associated with OS, a multivariable Cox proportional hazards regression model was fitted among patients with metastatic disease. The model was stratified by cancer type to allow for cancer type-specific baseline hazards. Adjusted hazard ratios (HRs) with 95% CIs were reported.

To contextualize real-world outcomes, pivotal phase III randomized controlled trials were identified by cancer type and treatment regimen, focusing on combinations known to represent first-line treatment in routine clinical practice. Published Kaplan–Meier survival estimates were digitized using WebPlotDigitizer and plotted alongside FAIR-ICI estimates for descriptive comparison.

The primary descriptive analyses and dashboard-based subgroup exploration were planned, while comparisons with pivotal clinical trials and post-discontinuation survival analyses were added during the study.

### Interactive dashboard

To facilitate transparent exploration of the FAIR-ICI cohort, we developed an interactive dashboard that provides descriptive summaries of patient characteristics, treatment patterns, and clinical outcomes, with dynamic filtering by tumor type, treatment regimen, demographic characteristics, and disease features. Users can define granular subgroups of interest (e.g., women aged >70 years with metastatic breast cancer treated with anti-PD-1) and explore corresponding treatment trajectories and outcome estimates. The dashboard is publicly available at [FAIR-ICI dashboard link](https://app.powerbi.com/view?r=eyJrIjoiMjQ5ZDczYjQtOThlMS00ZGM3LThlMzEtODE4ZDQxMTg0NWQ4IiwidCI6Ijk3MjYyNjI0LTZhOTctNDFjZS04NjAyLTIyZmM0M2I3MzYzYSIsImMiOjl9). Small cells are masked.

## Results

### Patient cohort

Of 3,780 patients with recorded ICI exposure, 3,231 met the eligibility criteria and were included in the final FAIR-ICI cohort (details in **Figure 1**). The cohort included patients receiving anti-PD-1, anti-PD-L1, and anti-CTLA-4 therapies, either alone or in combination.

**Figure 1.**
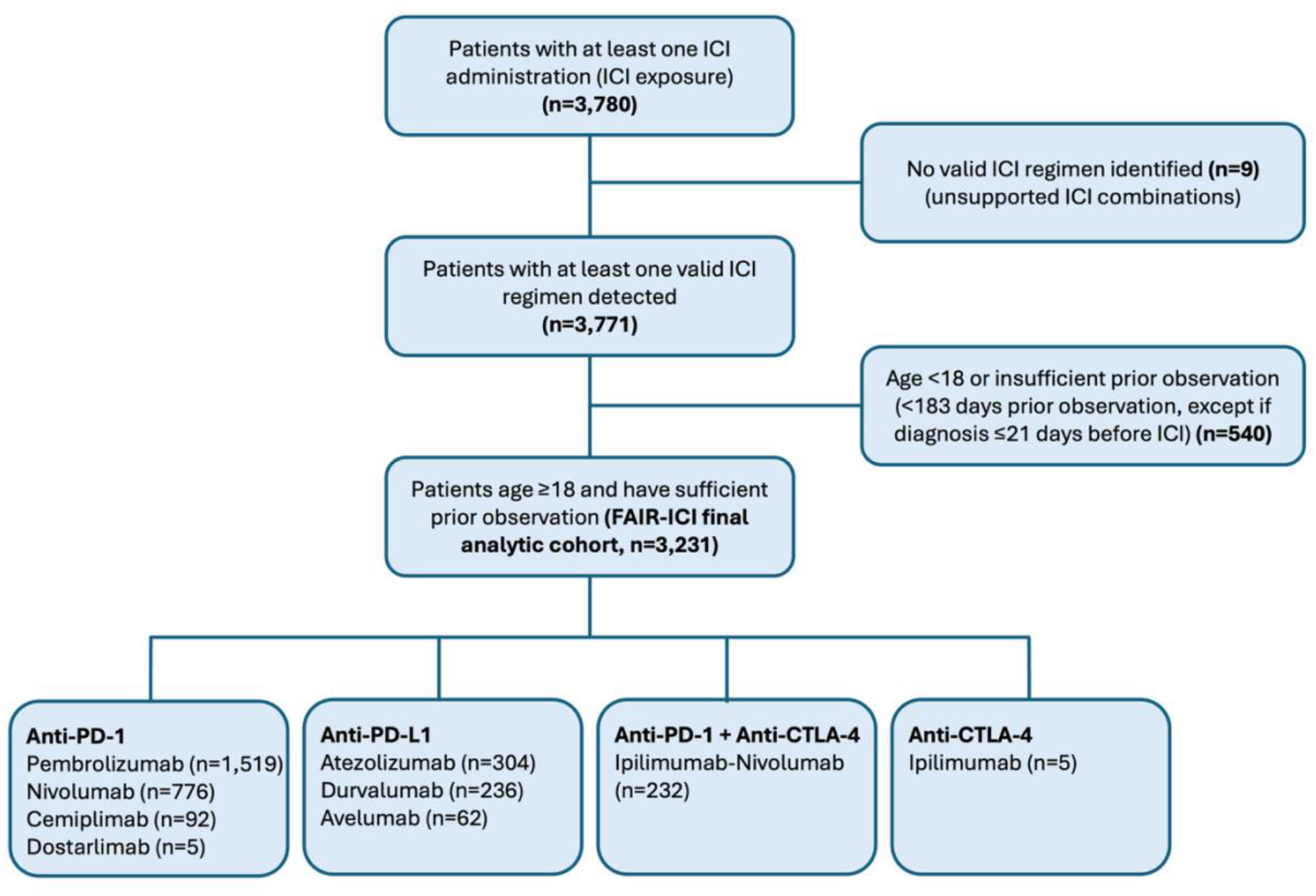
FAIR-ICI study cohort selection. A valid ICI regimen was defined as an ICI treatment regimen recognized by the predefined study algorithm, which recognized ICI monotherapy and nivolumab–ipilimumab dual therapy. Unsupported ICI combinations were excluded. **Abbreviations**: ICI, immune checkpoint inhibitor; PD-1, programmed cell death protein 1; PD-L1, programmed death-ligand 1; CTLA-4, cytotoxic T-lymphocyte-associated protein 4.

### Baseline characteristics

Overall, the cohort was predominantly older (59.7% aged ≥65 years) and male (62.5%) (**Table 1**). ICI uptake increased over the years, from 185 patients during 2015–2017 to 598 in 2022. Most patients had metastatic disease at treatment initiation (77.8%), but the number of patients receiving ICI in the non-metastatic setting increased markedly over time, from 5 patients (1.5% of ICI initiations) in 2018 to 71 (12.3%) in 2023. Treatment patterns varied across cancer types. ICI monotherapy was the most common regimen (55.0%), particularly in melanoma and urothelial carcinoma, whereas ICI combined with chemotherapy was more frequent in NSCLC.

**Table 1.** Baseline characteristics at ICI initiation, stratified by tumor type.

| Characteristic | Overall | NSCLC | Melanoma | Head and neck | Urothelial | RCC | Others, known * | Unknown** |
|---|---|---|---|---|---|---|---|---|
| No. | 3231 | 1007 | 358 | 258 | 221 | 207 | 894 | 286 |
| **Demographics** |  |  |  |  |  |  |  |  |
| **Age** |  |  |  |  |  |  |  |  |
| 18–49 | 285 (8.8%) | 49 (4.9%) | 50 (14.0%) | 32 (12.4%) | 6 (2.7%) | 11 (5.3%) | 108 (12.1%) | 29 (10.1%) |
| 50–64 | 1015 (31.4%) | 337 (33.5%) | 122 (34.1%) | 115 (44.6%) | 43 (19.5%) | 60 (29.0%) | 271 (30.3%) | 67 (23.4%) |
| 65–74 | 1125 (34.8%) | 404 (40.1%) | 88 (24.6%) | 69 (26.7%) | 83 (37.6%) | 75 (36.2%) | 312 (34.9%) | 94 (32.9%) |
| ≥75 | 806 (24.9%) | 217 (21.5%) | 98 (27.4%) | 42 (16.3%) | 89 (40.3%) | 61 (29.5%) | 203 (22.7%) | 96 (33.6%) |
| **Sex** |  |  |  |  |  |  |  |  |
| Female | 1212 (37.5%) | 345 (34.3%) | 161 (45.0%) | 62 (24.0%) | 50 (22.6%) | 47 (22.7%) | 434 (48.5%) | 113 (39.5%) |
| Male | 2019 (62.5%) | 662 (65.7%) | 197 (55.0%) | 196 (76.0%) | 171 (77.4%) | 160 (77.3%) | 460 (51.5%) | 173 (60.5%) |
| **BMI category** |  |  |  |  |  |  |  |  |
| Underweight (<18.5) | 185 (5.7%) | 54 (5.4%) | 5 (1.4%) | 46 (17.8%) | 10 (4.5%) | - | 49 (5.5%) | 17 (5.9%) |
| Normal (18.5–24.9) | 1026 (31.8%) | 352 (35.0%) | 80 (22.3%) | 109 (42.2%) | 52 (23.5%) | 52 (25.1%) | 291 (32.6%) | 90 (31.5%) |
| Overweight (25–29.9) | 894 (27.7%) | 281 (27.9%) | 110 (30.7%) | 54 (20.9%) | 62 (28.1%) | 63 (30.4%) | 236 (26.4%) | 88 (30.8%) |
| Obese (≥30) | 548 (17.0%) | 147 (14.6%) | 89 (24.9%) | 22 (8.5%) | 42 (19.0%) | 53 (25.6%) | 159 (17.8%) | 36 (12.6%) |
| Unknown | 578 (17.9%) | 173 (17.2%) | 74 (20.7%) | 27 (10.5%) | 55 (24.9%) | 35 (16.9%) | 159 (17.8%) | 55 (19.2%) |
| **Clinical / Lifestyle Characteristics** |  |  |  |  |  |  |  |  |
| **Performance status (ECOG)** |  |  |  |  |  |  |  |  |
| 0-1 | 1009 (31.2%) | 302 (30.0%) | 99 (27.7%) | 91 (35.3%) | 87 (39.4%) | 85 (41.1%) | 253 (28.3%) | 92 (32.2%) |
| ≥2 | 316 (9.8%) | 115 (11.4%) | 10 (2.8%) | 36 (14.0%) | 30 (13.6%) | 23 (11.1%) | 83 (9.3%) | 19 (6.6%) |
| Unknown | 1906 (59.0%) | 590 (58.6%) | 249 (69.6%) | 131 (50.8%) | 104 (47.1%) | 99 (47.8%) | 558 (62.4%) | 175 (61.2%) |
| **Smoking status** |  |  |  |  |  |  |  |  |
| Non-smoker | 449 (13.9%) | 63 (6.3%) | 57 (15.9%) | 32 (12.4%) | 38 (17.2%) | 43 (20.8%) | 166 (18.6%) | 50 (17.5%) |
| Smoker | 1050 (32.5%) | 477 (47.4%) | 34 (9.5%) | 106 (41.1%) | 62 (28.1%) | 44 (21.3%) | 256 (28.6%) | 71 (24.8%) |
| Ex-smoker | 539 (16.7%) | 246 (24.4%) | 29 (8.1%) | 48 (18.6%) | 36 (16.3%) | 26 (12.6%) | 116 (13.0%) | 38 (13.3%) |
| Unknown | 1193 (36.9%) | 221 (21.9%) | 238 (66.5%) | 72 (27.9%) | 85 (38.5%) | 94 (45.4%) | 356 (39.8%) | 127 (44.4%) |
| **Alcohol abuse** |  |  |  |  |  |  |  |  |
| Yes | 394 (12.2%) | 137 (13.6%) | 9 (2.5%) | 82 (31.8%) | 21 (9.5%) | 11 (5.3%) | 103 (11.5%) | 31 (10.8%) |
| No/Unknown | 2837 (87.8%) | 870 (86.4%) | 349 (97.5%) | 176 (68.2%) | 200 (90.5%) | 196 (94.7%) | 791 (88.5%) | 255 (89.2%) |
| **Disease characteristics at ICI start** |  |  |  |  |  |  |  |  |
| **Metastatic status** |  |  |  |  |  |  |  |  |
| Metastatic | 2514 (77.8%) | 897 (89.1%) | 297 (83.0%) | 215 (83.3%) | 182 (82.4%) | 190 (91.8%) | 733 (82.0%) | - |
| Non-metastatic | 197 (6.1%) | 58 (5.8%) | 26 (7.3%) | <5 | 13 (5.9%) | 11 (5.3%) | 86 (9.6%) | - |
| Inconclusive | 520 (16.1%) | 52 (5.2%) | 35 (9.8%) | 40 (15.5%) | 26 (11.8%) | 6 (2.9%) | 75 (8.4%) | 286 (100%) |
| **Stage** |  |  |  |  |  |  |  |  |
| Stage I | 37 (1.1%) | 5 (0.5%) | <5 | - | <5 | <5 | 21 (1.8%) | - |
| Stage II | 88 (2.7%) | 7 (0.7%) | 13 (3.6%) | 5 (1.9%) | 6 (2.7%) | - | 57 (4.8%) | - |
| Stage III | 158 (4.9%) | 62 (6.2%) | 19 (5.3%) | <5 | 6 (2.7%) | 12 (5.8%) | 56 (4.7%) | - |
| Stage IV | 2465 (76.3%) | 909 (90.3%) | 298 (83.2%) | 210 (81.4%) | 181 (81.9%) | 189 (91.3%) | 678 (57.5%) | - |
| Unknown | 483 (14.9%) | 24 (2.4%) | 24 (6.7%) | 40 (15.5%) | 24 (10.9%) | <5 | 82 (9.2%) | 286 (100%) |
| **Treatment details** |  |  |  |  |  |  |  |  |
| **ICI start year** |  |  |  |  |  |  |  |  |
| 2015–2017 | 185 (5.7%) | 56 (5.6%) | 53 (14.8%) | 18 (7.0%) | 16 (7.2%) | 15 (7.2%) | 18 (2.0%) | 9 (3.1%) |
| 2018 | 327 (10.1%) | 141 (14.0%) | 35 (9.8%) | 26 (10.1%) | 19 (8.6%) | 14 (6.8%) | 67 (7.5%) | 25 (8.7%) |
| 2019 | 404 (12.5%) | 144 (14.3%) | 43 (12.0%) | 37 (14.3%) | 32 (14.5%) | 35 (16.9%) | 75 (8.4%) | 38 (13.3%) |
| 2020 | 396 (12.3%) | 137 (13.6%) | 63 (17.6%) | 33 (12.8%) | 30 (13.6%) | 24 (11.6%) | 75 (8.4%) | 34 (11.9%) |
| 2021 | 458 (14.2%) | 162 (16.1%) | 49 (13.7%) | 44 (17.1%) | 31 (14.0%) | 31 (15.0%) | 100 (11.2%) | 41 (14.3%) |
| 2022 | 598 (18.5%) | 172 (17.1%) | 51 (14.2%) | 42 (16.3%) | 41 (18.6%) | 43 (20.8%) | 204 (22.8%) | 45 (15.7%) |
| 2023*** | 578 (17.9%) | 135 (13.4%) | 37 (10.3%) | 40 (15.5%) | 35 (15.8%) | 35 (16.9%) | 231 (25.8%) | 65 (22.7%) |
| 2024*** | 285 (8.8%) | 60 (6.0%) | 27 (7.5%) | 18 (7.0%) | 17 (7.7%) | 10 (4.8%) | 124 (13.9%) | 29 (10.1%) |
| **ICI mechanisms** |  |  |  |  |  |  |  |  |
| **Anti-PD-1** |  |  |  |  |  |  |  |  |
| Pembrolizumab | 1519 (47.0%) | 706 (70.1%) | 87 (24.3%) | 126 (48.8%) | 128 (57.9%) | 50 (24.2%) | 330 (36.9%) | 92 (32.2%) |
| Nivolumab | 776 (24.0%) | 102 (10.1%) | 181 (50.6%) | 132 (51.2%) | 28 (12.7%) | 96 (46.4%) | 125 (14.0%) | 112 (39.2%) |
| Cemiplimab | 92 (2.8%) | 13 (1.3%) | - | - | - | - | 55 (6.2%) | 24 (8.4%) |
| Dostarlimab | 5 (0.2%) | - | - | - | - | - | <5 | <5 |
| **Anti-PD-L1** |  |  |  |  |  |  |  |  |
| Atezolizumab | 304 (9.4%) | 71 (7.1%) | - | - | 24 (10.9%) | - | 187 (20.9%) | 22 (7.7%) |
| Durvalumab | 236 (7.3%) | 90 (8.9%) | - | - | - | - | 124 (13.9%) | 22 (7.7%) |
| Avelumab | 62 (1.9%) | - | - | - | 41 (18.6%) | <5 | 14 (1.6%) | 6 (2.1%) |
| **Anti-CTLA-4** |  |  |  |  |  |  |  |  |
| Ipilimumab | 5 (0.2%) | - | 5 (1.4%) | - | - | - | - | - |
| **Anti-PD-1 + Anti-CTLA-4** |  |  |  |  |  |  |  |  |
| Ipilimumab-Nivolumab | 232 (7.2%) | 25 (2.5%) | 85 (23.7%) | - | - | 60 (29.0%) | 55 (6.2%) | 7 (2.4%) |
| **Treatment regimen group** |  |  |  |  |  |  |  |  |
| ICI mono | 1776 (55.0%) | 518 (51.4%) | 258 (72.1%) | 179 (69.4%) | 200 (90.5%) | 91 (43.9%) | 347 (38.8%) | 183 (64.0%) |
| ICI dual (+ others) | 240 (7.4%) | 26 (2.6%) | 88 (24.6%) | <5 | - | 61 (29.5%) | 55 (6.2%) | 7 (2.4%) |
| ICI + chemo | 1005 (31.1%) | 450 (44.7%) | <5 | 61(23.6%) | 15 (6.8%) | <5 | 410 (45.9%) | 62 (21.7%) |
| ICI + TKI | 91 (2.8%) | 6 (0.6%) | 7 (1.9%) | - | - | 35 (16.9%) | 26 (2.9%) | 17 (5.9%) |
| ICI + other | 119 (3.7%) | 7 (0.7%) | <5 | 15 (5.8%) | 6 (2.7%) | 17 (8.2%) | 56 (6.3%) | 17 (5.9%) |

\*Including lung cancer (unspecified), breast cancer, small cell lung cancer (SCLC), mesothelioma, hepatocellular carcinoma (HCC), biliary tract cancer, cervical cancer, colorectal cancer, gastric cancer, endometrial cancer, cutaneous squamous cell carcinoma, esophageal adenocarcinoma, Merkel cell carcinoma, esophageal squamous cell carcinoma, basal cell carcinoma, Hodgkin lymphoma and gallbladder carcinoma. Details are shown in Supplementary Table S5 and can also be explored in the FAIR-ICI dashboard.
\*\*Non-reimbursed indications in Belgium at the time of analysis (e.g. compassionate use).
\*\*\*Incomplete data for 2023 and 2024 because the patient identification period ended before the end of the calendar year to allow sufficient follow-up.
**Abbreviations:** BMI, body mass index; ECOG, Eastern Cooperative Oncology Group; ICI, immune checkpoint inhibitor; NSCLC, non-small cell lung cancer; RCC, renal cell carcinoma; chemo, chemotherapy; TKI, tyrosine kinase inhibitor.

### Treatment discontinuation patterns and post-discontinuation survival

Among metastatic patients (**Figure 2A**), the majority (62%, n=1,548) either died during treatment (23.9%, n=601) or initiated a new regimen within 12 weeks of the last ICI administration (37.7%, n=947), the latter potentially reflecting progression or toxicity. In contrast, 22.6% (n=568) discontinued ICI without subsequent treatment within 12 weeks, potentially reflecting deep remission or toxicity without the need or possibility for further treatment in the short term. This was most frequent in melanoma (104/297, 35.0%) and RCC (39/190, 20.5%), consistent with known ICI efficacy, and least frequent in head and neck cancer (33/215, 15.3%).

**Figure 2.**
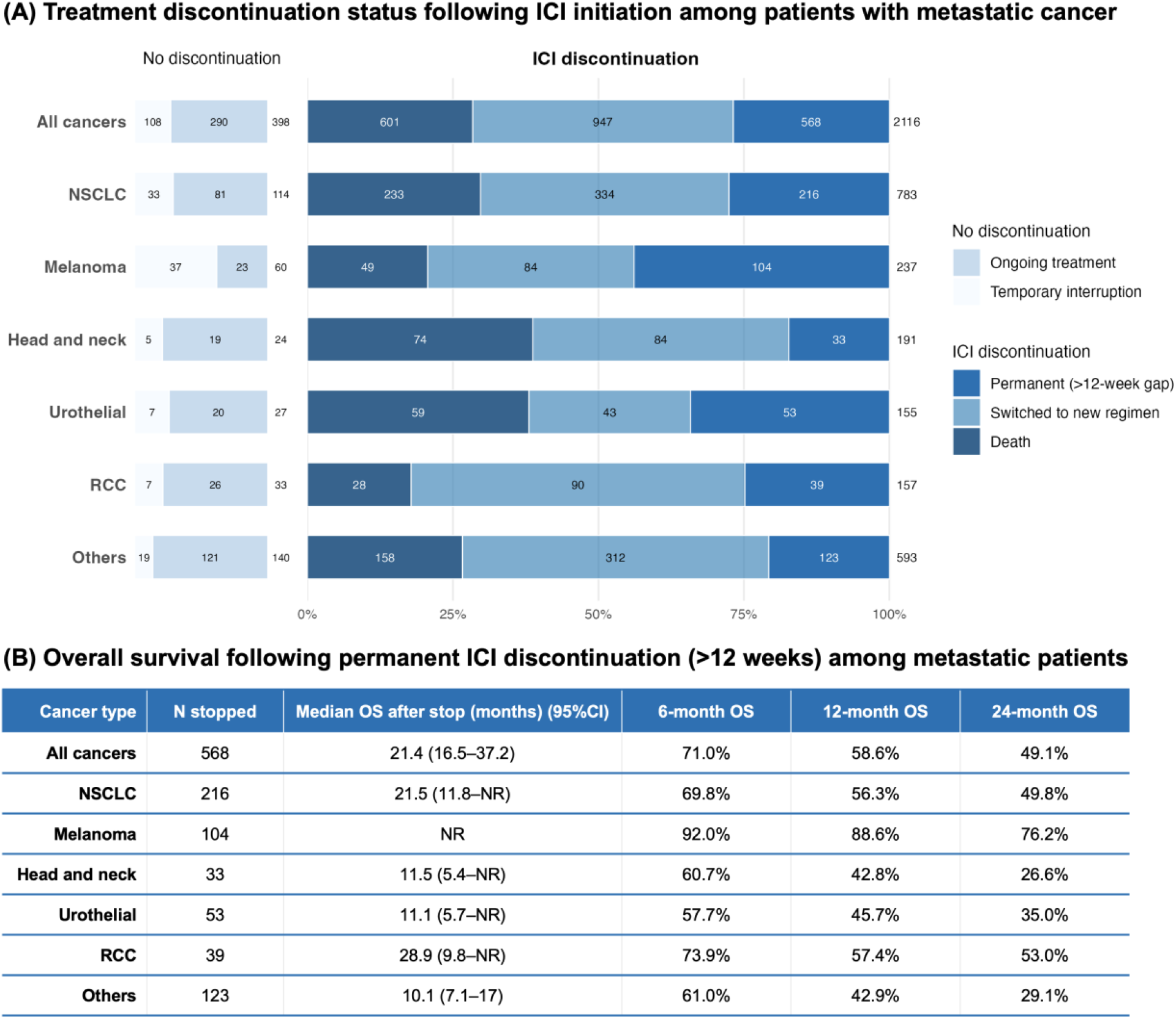
Treatment discontinuation patterns and overall survival following permanent ICI discontinuation among metastatic patients receiving ICIs (n=2,514). Permanent ICI discontinuation was defined as no ICI administration for >12 weeks after the last ICI administration. Patients who subsequently re-initiated ICI after this 12-week gap were classified separately as “temporary interruption”. Overall survival was estimated from the date of permanent ICI discontinuation (12 weeks after the last ICI administration). **Abbreviations**: CI, confidence interval; ICI, immune checkpoint inhibitor; OS, overall survival; NSCLC, non-small cell lung cancer; RCC, renal cell carcinoma.

Among patients permanently discontinuing ICI without subsequent treatment (**Figure 2B**), median post-discontinuation OS was 21.4 months (95% CI 16.5–37.2). Post-discontinuation survival was most favorable in melanoma (median not reached; 24-month OS 76.2%), followed by RCC (median 28.9 months, 95% CI 9.8–NR; 24-month OS 53.0%). Corresponding 24-month OS estimates were 49.8% for NSCLC, 35.0% for urothelial carcinoma, and 26.6% for head and neck cancer.

### Real-world survival and treatment outcomes

Among metastatic patients, median OS varied substantially across the five major cancer types, ranging from 11 months in head and neck cancer and urothelial carcinoma to 78 months in melanoma (**Table 2**). Twelve-month OS ranged from 44.1% to 75.4%, while 24-month OS ranged from 24.9% to 67.2%.

**Table 2.** Real-world survival and treatment outcomes by cancer type in metastatic patients receiving ICIs.

| **Cancer type** | **n** | **Outcome** | **Median (months)<br>(95%CI)** | **12-month<br>KM estimate (%)** | **24-month<br>KM estimate (%)** |
|---|---|---|---|---|---|
| NSCLC | 897 | OS | 15 (13–18) | 55.1 | 39.3 |
|  |  | ToT | 7 (6–7) | 31.6 | 16.0 |
|  |  | TTNT | 14 (11–18) | 51.6 | 41.7 |
| Melanoma | 297 | OS | 78 (40–NA) | 72.2 | 67.2 |
|  |  | ToT | 7 (6–11) | 38.0 | 10.4 |
|  |  | TTNT | 26 (16–56) | 63.1 | 51.9 |
| Head and neck | 215 | OS | 11 (8–14) | 44.2 | 24.9 |
|  |  | ToT | 4 (4–6) | 17.5 | 7.1 |
|  |  | TTNT | 8 (6–14) | 42.1 | 33.3 |
| Urothelial | 182 | OS | 11 (8–15) | 44.1 | 28.9 |
|  |  | ToT | 6 (5–9) | 32.8 | 20.4 |
|  |  | TTNT | 25 (14–57) | 62.0 | 50.4 |
| RCC | 190 | OS | 33 (24–38) | 75.4 | 55.9 |
|  |  | ToT | 6 (5–9) | 30.9 | 14.0 |
|  |  | TTNT | 11 (8–15) | 48.3 | 28.8 |

Other cancer types are presented in Supplementary Table S7. Kaplan–Meier curves for all outcomes and cancer types can be explored in the FAIR-ICI dashboard.
**Abbreviations**: CI, confidence interval; ICI, immune checkpoint inhibitor; NSCLC, non-small cell lung cancer; RCC, renal cell carcinoma; OS, overall survival; ToT, time on treatment; TTNT, time to next treatment. Median OS, ToT, and TTNT were estimated using the Kaplan–Meier method. Twelve- and 24-month estimates represent Kaplan–Meier estimates at the corresponding time points.

Among these five cancer types, OS event proportions ranged from 32.7% in melanoma to 66.0% in head and neck cancer, while median follow-up ranged from 7.6 months in head and neck cancer to 20.8 months in melanoma. Detailed maturity metrics, including follow-up among surviving patients and the number of ICI administrations, are presented in **Supplementary-Table S6**.

Median ToT was consistently shorter than median OS, ranging from 4 to 7 months, corresponding to a median of 5–6 ICI administrations (Supplementary-Table S6). TTNT showed greater variation, ranging from 8 months in head and neck cancer to 26 months in melanoma. This discrepancy between treatment duration and survival was particularly pronounced in melanoma, where median OS reached 78 months despite a median ToT of 7 months and median TTNT of 26 months.

### Comparison with pivotal clinical trials

We compared real-world outcomes in unselected patient populations with pivotal clinical trials for all cancer type–regimen subgroups known to represent first-line treatment settings (**Figure 3**). Real-world survival curves generally followed patterns similar to the corresponding pivotal clinical trials but were frequently less favorable, particularly during early follow-up. The most pronounced visual discrepancies were observed in NSCLC treated with pembrolizumab plus chemotherapy, HCC with atezolizumab plus bevacizumab, and RCC with pembrolizumab plus TKI. In contrast, closer alignment was observed for RCC and mesothelioma treated with ipilimumab plus nivolumab, and head and neck cancer treated with pembrolizumab plus chemotherapy.

**Figure 3.**
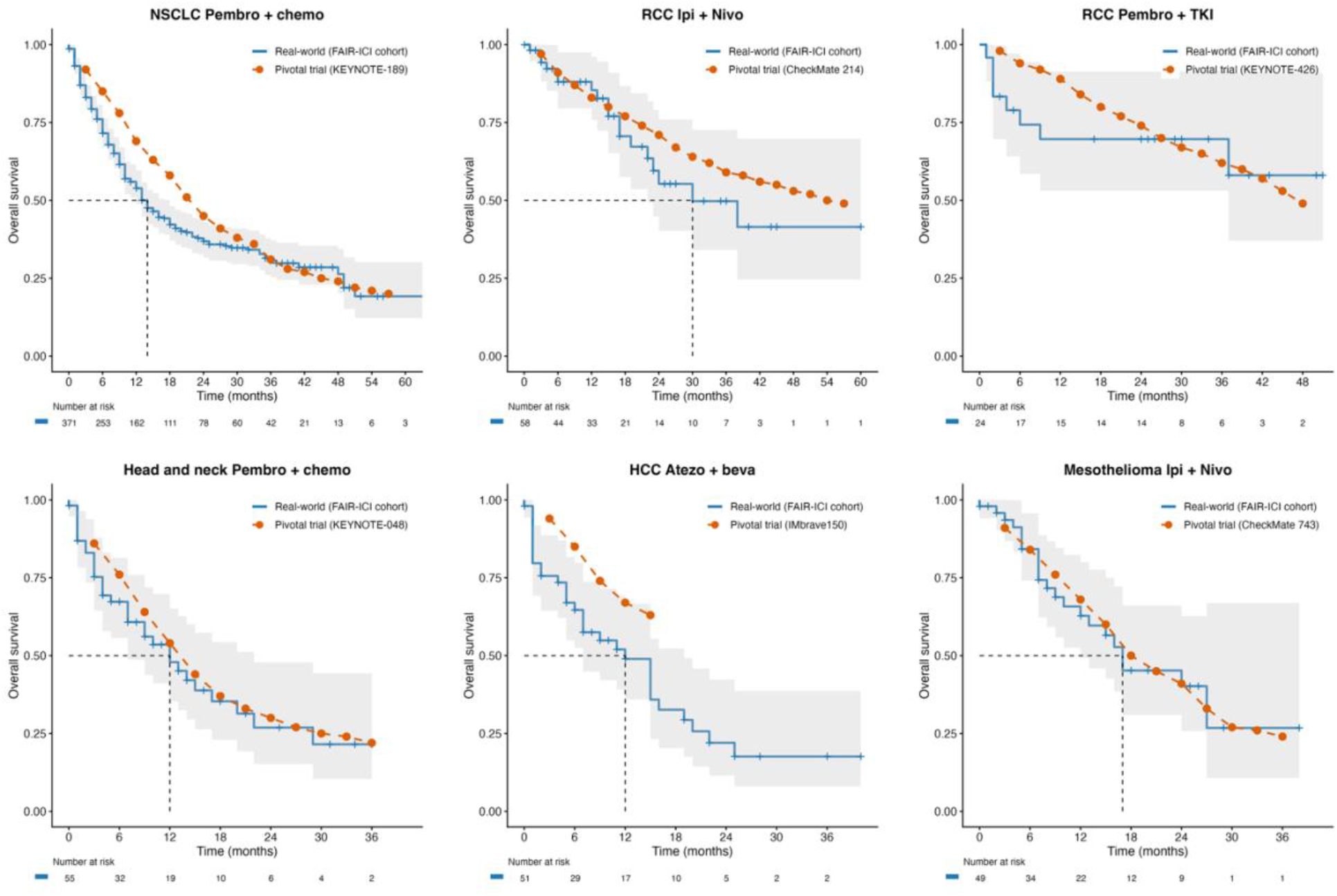
Real-world overall survival compared with pivotal clinical trials across selected cancer types and regimens. Trial survival curves were digitized from published Kaplan–Meier plots of the corresponding pivotal clinical trial, using the most recent available follow-up where available. **Abbreviations**: OS, overall survival; NSCLC, non-small cell lung cancer; RCC, renal cell carcinoma; HCC, hepatocellular carcinoma; ICI, immune checkpoint inhibitor; TKI, tyrosine kinase inhibitor; chemo, chemotherapy; Pembro, pembrolizumab; Atezo, atezolizumab; Nivo, nivolumab; Ipi, ipilimumab; Beva, bevacizumab.

### Survival according to ECOG performance status

Patients with ECOG 0–1 consistently showed longer OS than those with ECOG ≥2 (**Figure 4**), with significant differences in NSCLC, melanoma, RCC, and urothelial carcinoma, but not head and neck cancer. Among patients with NSCLC receiving first-line pembrolizumab plus chemotherapy, survival in patients with ECOG 0–1 more closely approximated that reported in the pivotal KEYNOTE-189 trial, whereas patients with ECOG ≥2 had substantially poorer survival.

**Figure 4.**
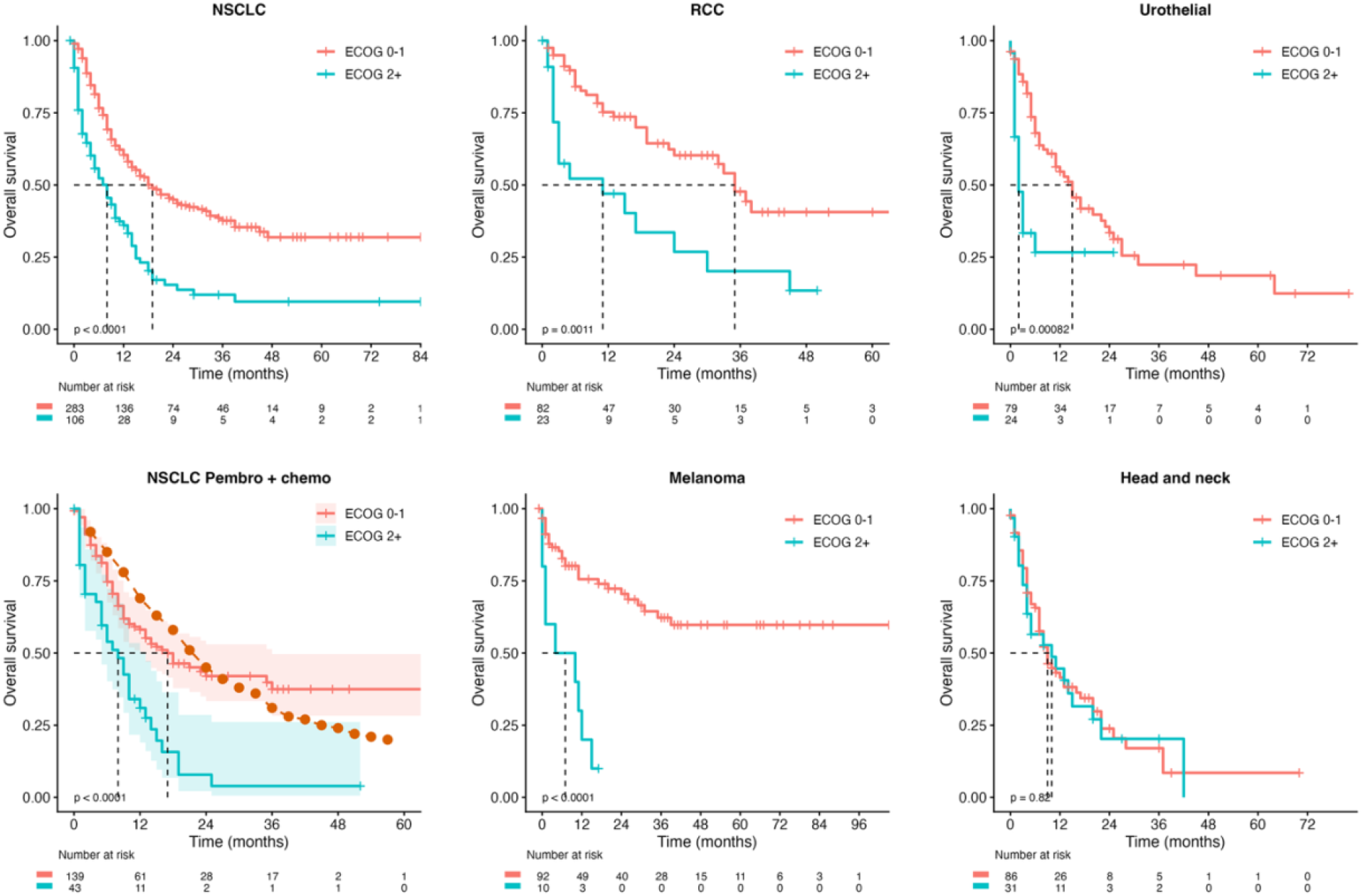
Real-world overall survival stratified by ECOG performance status among metastatic patients receiving ICIs, by cancer type. In the NSCLC pembrolizumab plus chemotherapy panel, the orange dotted line represents overall survival reported in the pivotal KEYNOTE-189 trial, using the most recent available follow-up. **Abbreviations**: ECOG, Eastern Cooperative Oncology Group; ICI, immune checkpoint inhibitor; NSCLC, non-small cell lung cancer; RCC, renal cell carcinoma; chemo, chemotherapy; Pembro, pembrolizumab.

### Factors associated with overall survival

Additional Kaplan-Meier analyses by age, sex, BMI, smoking status, and alcohol abuse are presented in **Supplementary-Figures S3–S7**. Modest differences in survival by age and sex were observed in selected cancers, particularly melanoma and RCC, although these patterns were not consistently significant across tumor types. In contrast, associations with BMI, smoking status, and alcohol abuse were less consistent.

In multivariable Cox regression among metastatic patients (**Table 3**), older age, male sex, and ECOG ≥2 were associated with poorer OS. ECOG ≥2 showed the strongest association with mortality (adjusted HR 2.18, 95% CI 1.82–2.60), while mortality increased progressively with age. No clear associations were observed for BMI, smoking status, or alcohol abuse.

**Table 3.**
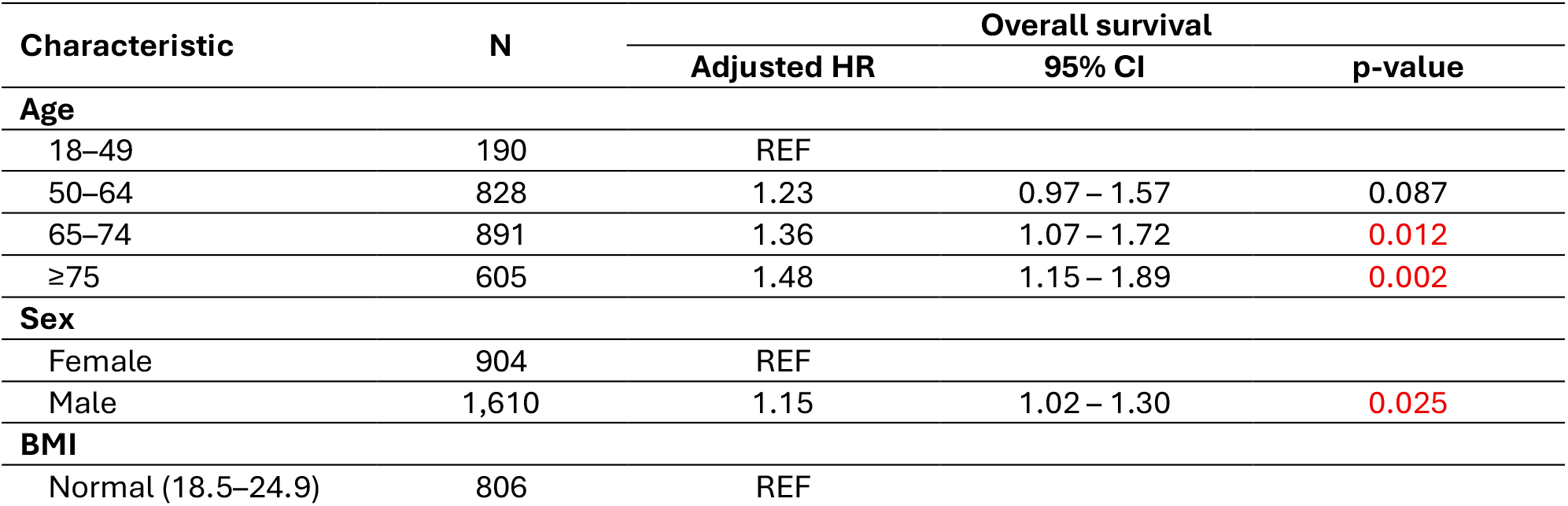
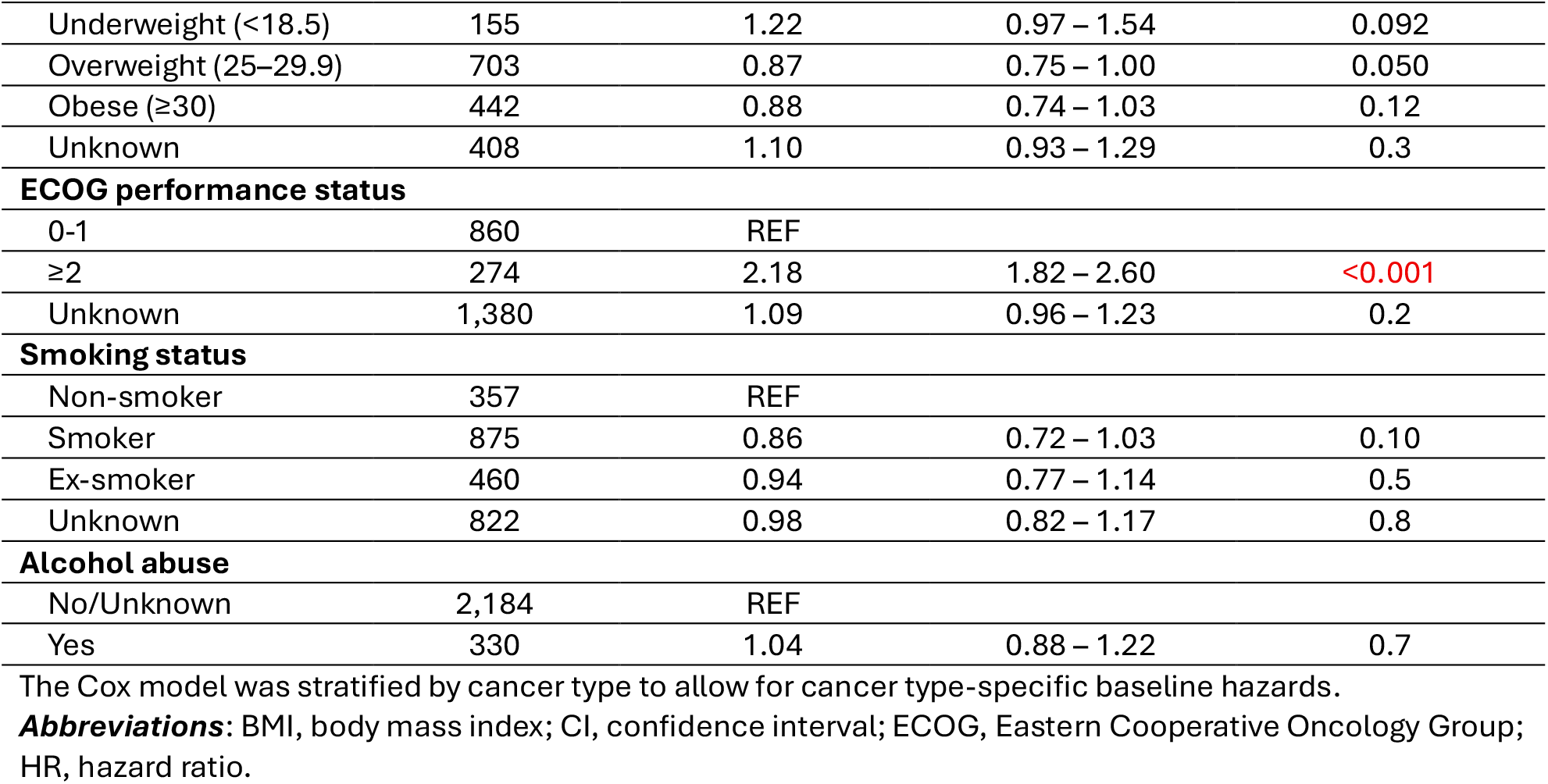
Multivariable Cox proportional hazards analysis of overall survival among metastatic patients receiving ICIs (n=2,514)

## Discussion

In this multicenter real-world cohort of 3,231 patients receiving ICIs across four Belgian hospitals, we described treatment utilization, trajectories, and clinical outcomes across multiple cancer types using an NLP-enriched OMOP-CDM framework. Several key findings warrant particular attention. First, nearly one-quarter of metastatic patients permanently discontinued ICIs without initiating another systemic therapy within 12 weeks and nevertheless experienced prolonged survival (2-year OS 49% after stop). Second, treatment-based endpoints (ToT and TTNT) were substantially shorter than OS across tumor types, suggesting that many patients remained alive long after treatment discontinuation and often before requiring additional therapy. Third, real-world outcomes generally followed pivotal trial pattern but were frequently less favorable, emphasizing the need for complementary RWE alongside RCTs. Finally, ECOG performance status emerged as one of the strongest prognostic factors across analyses, highlighting the importance of systematic assessment in routine oncology practice.

### Treatment discontinuation and durable benefit

One of the most notable findings was that permanent ICI discontinuation did not necessarily indicate treatment failure or poor prognosis (**Figure 2**). Although discontinuation reasons require further validation, preliminary analyses suggest a heterogeneous mixture of treatment-related toxicity, disease progression, patient preference, and sustained disease control without immediate need for subsequent systemic therapy. Favorable post-discontinuation outcomes, particularly in melanoma and RCC, suggest durable benefit beyond active treatment in a subset of patients.

These observations are consistent with growing evidence that durable disease control can persist after treatment cessation. ICIs may promote durable antitumor immune memory, sustaining T-cell-mediated tumor control beyond active treatment. Optimal duration of ICI treatment remains uncertain, with pivotal trials generally continuing treatment until progression, unacceptable toxicity, or a maximum of two years. In melanoma, long-term trial evidence has demonstrated durable disease control and survival following ICI therapy^18,19^. RWE further showed low relapse after elective anti-PD-1 discontinuation among patients achieving complete response after more than six months of treatment^20^, while a recent emulated target trial suggested that around one year may be sufficient in patients with disease control^21^. Similarly, in metastatic RCC, elective discontinuation after approximately two years did not appear to compromise long-term outcomes among responding patients ^22^. Collectively, these findings suggest that cessation does not necessarily indicate loss of clinical benefit and highlight the need for further investigation of optimal ICI duration in routine practice^23^.

The discrepancy observed between OS and treatment-trajectory endpoints further supports this interpretation. Across tumor types, median ToT and TTNT were consistently shorter than OS (**Table 2**), indicating that many patients remained alive for prolonged periods after treatment discontinuation and often before requiring subsequent systemic therapy. These findings suggest that treatment duration, switching, and survival capture distinct aspects of real-world treatment trajectories. Consequently, ToT and TTNT should be considered complementary treatment-trajectory endpoints that provide information distinct from OS, such as health care consumption, when evaluating real-world immunotherapy outcomes^12,24^.

### Real-world versus trial overall survival

Compared with pivotal clinical trials, real-world survival was frequently less favorable, particularly during early follow-up. The most pronounced visual discrepancies were observed in NSCLC treated with pembrolizumab plus chemotherapy, HCC with atezolizumab plus bevacizumab, and RCC with pembrolizumab plus TKI (**Figure 3**). These findings are consistent with the well-recognized differences between trial populations and routine clinical practice. Pivotal trials often employ restrictive eligibility criteria that exclude patients with poor performance status, substantial comorbidities, organ dysfunction, autoimmune disease, or advanced age, thereby limiting the representativeness of enrolled populations^9,10,25^. Recent analyses also identified prognostic heterogeneity as an important explanation, with high-risk real-world patients experiencing substantially poorer survival than would be predicted from pivotal trials^26^. Our ECOG-stratified analysis of NSCLC patients receiving pembrolizumab plus chemotherapy further illustrates this (**Figure 4**). Survival with ECOG 0–1 more closely approximated KEYNOTE-189, whereas patients with ECOG ≥2 had substantially poorer survival, suggesting that performance status contributes to trial–real-world differences.

The observed RCC treatment patterns further highlight the complexity of interpreting real-world comparative effectiveness. While outcomes with nivolumab plus ipilimumab were broadly comparable to CheckMate214, early survival with pembrolizumab plus TKI appeared less favorable than in KEYNOTE-426. Although the number of patients receiving pembrolizumab plus TKI was small (n=24) and no comparative conclusions can be drawn, one possible explanation is confounding by indication. In routine practice, ICI–TKI combinations may be preferentially selected for patients with higher disease burden, more symptomatic disease, or a need for rapid disease control, given the high response rate (59.3%) and early tumor responses in metastatic RCC^27^. Consequently, patients receiving apparently similar first-line therapies in real-world settings may differ substantially in underlying prognosis, introducing biases that are difficult to fully capture using routinely collected data^26^. This underscores the importance of considering treatment-selection processes and baseline prognosis when interpreting real-world comparative effectiveness studies.

### Prognostic factors and the importance of ECOG performance status

Among the prognostic factors evaluated, ECOG performance status emerged as the strongest predictor of survival in both subgroup and multivariable analyses (**Figure 4**; **Table 3**). Patients with ECOG ≥2 had more than twice the hazard of death compared with ECOG 0–1, consistent with extensive literature identifying functional status as a major determinant of outcomes in patients receiving immunotherapy^28–31^. This is particularly relevant because pivotal trials predominantly enrolled patients with ECOG 0–1, whereas routine clinical practice includes a broader spectrum of patients, including those with impaired functional status. Consequently, performance status may contribute substantially to the differences observed between trial and real-world outcomes^9^.

Notably, ECOG performance status was unavailable for a substantial proportion of patients in our cohort (59%), reflecting a common limitation of routinely collected data. Nevertheless, the strong and consistent association observed among patients with available data suggests that performance status may explain an important component of outcome heterogeneity in routine practice. Given its strong prognostic value, more systematic capture of ECOG in EHRs should be prioritized to improve risk adjustment and interpretability of future real-world studies. The ability of FAIR-ICI to extract ECOG information through NLP-enhanced extraction from unstructured clinical documentation highlights the value of integrating structured and unstructured data sources for oncology RWE.

Beyond performance status, older age and male sex were associated with poorer OS in the multivariable analysis (**Table 3**), although differences were generally modest in cancer-specific Kaplan–Meier analyses (Supplementary-Figures S3–S4). Associations with BMI, smoking status, and alcohol abuse were less evident, with no clear associations observed after adjustment.

### Strengths and limitations

To our knowledge, FAIR-ICI represents the first multicenter immunotherapy-focused RWE platform integrating structured and unstructured EHR data within a standardized OMOP-CDM framework. By combining data from four hospitals and incorporating NLP-derived clinical variables, FAIR-ICI enabled evaluation of treatment trajectories and outcomes across multiple tumor types with greater clinical granularity than typically available in administrative or registry-based datasets. The reuse of routinely collected clinical data also enables large-scale analyses without additional burden to patients or resource-intensive prospective data collection. Furthermore, the publicly available interactive dashboard supports transparent exploration of results and facilitates subgroup analyses by clinicians, researchers, and policymakers.

Several limitations should be acknowledged. First, the retrospective observational design is inherently susceptible to residual confounding and treatment-selection bias. In particular, treatment allocation in routine clinical practice may be influenced by factors that were not fully captured in the available data, including disease burden, symptom severity, physician preference, and other clinical considerations, potentially resulting in confounding by indication. Second, several variables contained substantial missingness, particularly ECOG performance status and some lifestyle characteristics. Missing ECOG data may also introduce bias if their availability is associated with patient prognosis. Third, treatment line could not always be determined directly, and selected trial comparisons therefore relied on treatment-regimen subgroups considered likely to represent first-line settings. Fourth, reasons for treatment discontinuation were not directly assessed, limiting interpretation of the clinical circumstances underlying ICI cessation. Further characterization of discontinuation reasons and treatment-related toxicity represents an important direction for future FAIR-ICI analyses. Finally, despite including four hospitals, several cancer type–treatment subgroups remained small, limiting the precision of subgroup analyses and restricting interpretation. Belgium’s broad ICI access and the inclusion of academic and regional hospitals support the relevance of these findings to routine oncology practice. However, differences in reimbursement policies, patient characteristics, and data availability may limit generalizability to other healthcare systems.

## Conclusion

This multicenter real-world study provides a comprehensive overview of immunotherapy utilization, treatment trajectories, and outcomes across multiple cancer types. A notable proportion of patients remained alive for prolonged periods after treatment discontinuation without subsequent systemic therapy, highlighting the potential for durable clinical benefit beyond active treatment exposure. Differences between OS, ToT, and TTNT demonstrate that these endpoints capture distinct, complementary dimensions of immunotherapy benefit. While real-world outcomes broadly mirrored those observed in pivotal clinical trials, they were frequently less favorable, underscoring the importance of understanding differences between trial populations and routine practice. ECOG performance status was strongly associated with survival, emphasizing the importance of functional status in immunotherapy-treated populations and the need for its systematic capture in real-world datasets. The interactive dashboard further enables exploration of FAIR-ICI findings across clinically relevant patient subgroups. Together, these findings establish FAIR-ICI as a valuable RWE platform for future investigations, particularly for extending the dataset into treatment toxicity and reasons for treatment discontinuation, as well as treatment sequencing, and biomarker-driven precision immuno-oncology.

## Declarations

### Ethics approval and consent to participate

This study was approved by the Ethics Committees of the participating hospitals (reference UZA7026). Given the retrospective nature of the study, the requirement for individual informed consent was waived.

### Availability of data and material

The patient-level data used in this study are not publicly available because they contain sensitive health information and are subject to institutional and data governance restrictions. Aggregated results from the FAIR-ICI cohort can be explored through the publicly accessible interactive dashboard at [FAIR-ICI dashboard link](https://app.powerbi.com/view?r=eyJrIjoiMjQ5ZDczYjQtOThlMS00ZGM3LThlMzEtODE4ZDQxMTg0NWQ4IiwidCI6Ijk3MjYyNjI0LTZhOTctNDFjZS04NjAyLTIyZmM0M2I3MzYzYSIsImMiOjl9). Further information regarding access to the underlying data may be obtained from the corresponding author, subject to applicable ethical, institutional, and data-sharing requirements.

### Authors’ contributions

LTH: Conceptualization, Methodology, Formal analysis, Visualization, Investigation, Writing – original draft, Writing – review & editing.

FVD: Data curation, Methodology, Formal analysis, Software, Investigation, Writing – review & editing.

IB: Data curation, Methodology, Software, Investigation, Writing – review & editing.

CLO: Investigation, Writing – review & editing.

CV: Resources, Validation, Writing – review & editing.

DD: Validation, Writing – review & editing.

DH: Resources, Writing – review & editing. HP: Resources, Writing – review & editing.

LC: Validation, Writing – review & editing.

ADM: Validation, Writing – review & editing.

PRD: Resources, Validation, Writing – review & editing.

MV: Validation, Writing – review & editing.

SA: Resources, Validation, Supervision, Writing – review & editing. NB: Methodology, Supervision, Writing – review & editing.

AV: Conceptualization, Methodology, Supervision, Validation, Investigation, Funding acquisition, Resources, Writing – review & editing.

## Acknowledgements

The authors thank the IT and data teams at the participating hospitals for their technical support in data extraction and infrastructure. They also thank the EHDEN project for enabling the initial collaboration between hospitals and LynxCare to harmonize their health data to OMOP-CDM.

## Abbreviations

BMI: body mass index
CI: confidence interval
ECOG: Eastern Cooperative Oncology Group
EHR: electronic health record
EMA: European Medicines Agency
HCC: hepatocellular carcinoma
HR: hazard ratio
ICI: immune checkpoint inhibitor
IQR: interquartile range
NLP: natural language processing
NSCLC: non-small cell lung cancer
OMOP-CDM: Observational Medical Outcomes Partnership Common Data Model
OS: overall survival
RCC: renal cell carcinoma
RCT: randomized controlled trial
RWE: real-world evidence
TKI: tyrosine kinase inhibitor
ToT: time on treatment
TTNT: time to next treatment

## References

1. European Medicines Agency (EMA). European Public Assessment Report (EPAR): Yervoy. 2011. https://www.ema.europa.eu/en/medicines/human/EPAR/yervoy

2. Larroquette M, Domblides C, Lefort F, et al. Combining immune checkpoint inhibitors with chemotherapy in advanced solid tumours: A review. European Journal of Cancer. 2021;158:47–62. doi:10.1016/j.ejca.2021.09.013

3. Banna GL, Hassan MA, Signori A, et al. Neoadjuvant Chemo-Immunotherapy for Early-Stage Non–Small Cell Lung Cancer: A Systematic Review and Meta-Analysis. JAMA Netw Open. 2024;7(4):e246837. doi:10.1001/jamanetworkopen.2024.6837

4. Haslam A, Olivier T, Prasad V. How many people in the US are eligible for and respond to checkpoint inhibitors: An empirical analysis. International Journal of Cancer. 2025;156(12):2352–2359. doi:10.1002/ijc.35347

5. Belgian Cancer Registry. https://kankerregister.org/en

6. Maggie De Block. Immunotherapy reimbursement announcement. Maggie De Block. December 20, 2016. https://www.maggiedeblock.be/doeltreffender-behandeling-voor-10-000-kankerpatienten-immunotherapie-sneller-terugbetaald/

7. Belgian National Institute for Health and Disability Insurance (RIZIV). Accessed January 30, 2026. https://webappsa.riziv-inami.fgov.be/ssp/ProductSearch

8. Manzano A, Svedman C, Hofmarcher T, Wilking N. Comparator Report on Cancer in Europe 2025 - Disease Burden, Costs and Access to Medicines and Molecular Diagnostics. Published online 2025.

9. Kim ES, Bruinooge SS, Roberts S, et al. Broadening Eligibility Criteria to Make Clinical Trials More Representative: American Society of Clinical Oncology and Friends of Cancer Research Joint Research Statement. J Clin Oncol. 2017;35(33):3737–3744. doi:10.1200/JCO.2017.73.7916

10. Yoo SH, Keam B, Kim M, Kim TM, Kim DW, Heo DS. Generalization and representativeness of phase III immune checkpoint blockade trials in non-small cell lung cancer. Thoracic Cancer. 2018;9(6):736–744. doi:10.1111/1759-7714.12641

11. Khozin S, Blumenthal GM, Pazdur R. Real-world Data for Clinical Evidence Generation in Oncology. J Natl Cancer Inst. 2017;109(11). doi:10.1093/jnci/djx187

12. Stewart M, Norden AD, Dreyer N, et al. An Exploratory Analysis of Real-World End Points for Assessing Outcomes Among Immunotherapy-Treated Patients With Advanced Non–Small-Cell Lung Cancer. JCO Clin Cancer Inform. 2019;3:CCI.18.00155. doi:10.1200/CCI.18.00155

13. Di Maio M, Perrone F, Conte P. Real-World Evidence in Oncology: Opportunities and Limitations. Oncologist. 2020;25(5):e746–e752. doi:10.1634/theoncologist.2019-0647

14. Charlton ME, Kahl AR, McDowell BD, et al. Cancer Registry Data Linkage of Electronic Health Record Data From ASCO’s CancerLinQ: Evaluation of Advantages, Limitations, and Lessons Learned. JCO Clin Cancer Inform. 2022;(6):e2100149. doi:10.1200/CCI.21.00149

15. Hripcsak G, Duke JD, Shah NH, et al. Observational Health Data Sciences and Informatics (OHDSI): Opportunities for Observational Researchers. Stud Health Technol Inform. 2015;216:574–578.

16. Reich C, Ostropolets A, Ryan P, et al. OHDSI Standardized Vocabularies-a large-scale centralized reference ontology for international data harmonization. J Am Med Inform Assoc. 2024;31(3):583–590. doi:10.1093/jamia/ocad247

17. Van de Meulebroucke C, Beckers J, Corten K. What Can We Expect Following Anterior Total Hip Arthroplasty on a Regular Operating Table? A Validation Study of an Artificial Intelligence Algorithm to Monitor Adverse Events in a High-Volume, Nonacademic Setting. The Journal of Arthroplasty. 2019;34(10):2260–2266. doi:10.1016/j.arth.2019.07.039

18. Robert C, Ribas A, Schachter J, et al. Pembrolizumab versus ipilimumab in advanced melanoma (KEYNOTE-006): post-hoc 5-year results from an open-label, multicentre, randomised, controlled, phase 3 study. Lancet Oncol. 2019;20(9):1239–1251. doi:10.1016/S1470-2045(19)30388-2

19. Larkin J, Chiarion-Sileni V, Gonzalez R, et al. Five-Year Survival with Combined Nivolumab and Ipilimumab in Advanced Melanoma. New England Journal of Medicine. 2019;381(16):1535–1546. doi:10.1056/NEJMoa1910836

20. Jansen YJL, Rozeman EA, Mason R, et al. Discontinuation of anti-PD-1 antibody therapy in the absence of disease progression or treatment limiting toxicity: clinical outcomes in advanced melanoma. Ann Oncol. 2019;30(7):1154–1161. doi:10.1093/annonc/mdz110

21. Amiot M, Mortier L, Dalle S, et al. When to stop immunotherapy for advanced melanoma: the emulated target trials. eClinicalMedicine. 2024;78:102960. doi:10.1016/j.eclinm.2024.102960

22. Decruyenaere A, Christine G, Sylvie R, et al. Optimal treatment duration in metastatic renal cell carcinoma patients responding to immune checkpoint inhibitors: should we treat beyond two years? Acta Oncol. 2025;64:979–988. doi:10.2340/1651-226X.2025.43876

23. Tzeng A, Tzeng TH, Ornstein MC. Treatment-free survival after discontinuation of immune checkpoint inhibitors in metastatic renal cell carcinoma: a systematic review and meta-analysis. J Immunother Cancer. 2021;9(10):e003473. doi:10.1136/jitc-2021-003473

24. Blumenthal GM, Gong Y, Kehl K, et al. Analysis of time-to-treatment discontinuation of targeted therapy, immunotherapy, and chemotherapy in clinical trials of patients with non-small-cell lung cancer. Ann Oncol. 2019;30(5):830–838. doi:10.1093/annonc/mdz060

25. Singh H, Kanapuru B, Smith C, et al. FDA analysis of enrollment of older adults in clinical trials for cancer drug registration: A 10-year experience by the U.S. Food and Drug Administration. J Clin Oncol. 2017;35(15_suppl):10009–10009. doi:10.1200/JCO.2017.35.15_suppl.10009

26. Orcutt X, Chen K, Mamtani R, Long Q, Parikh RB. Evaluating generalizability of oncology trial results to real-world patients using machine learning-based trial emulations. Nat Med. 2025;31(2):457–465. doi:10.1038/s41591-024-03352-5

27. Rini BI, Plimack ER, Stus V, et al. Pembrolizumab plus Axitinib versus Sunitinib for Advanced Renal-Cell Carcinoma. New England Journal of Medicine. 2019;380(12):1116–1127. doi:10.1056/NEJMoa1816714

28. Facchinetti F, Mazzaschi G, Barbieri F, et al. First-line pembrolizumab in advanced non-small cell lung cancer patients with poor performance status. Eur J Cancer. 2020;130:155–167. doi:10.1016/j.ejca.2020.02.023

29. Dall’Olio FG, Maggio I, Massucci M, Mollica V, Fragomeno B, Ardizzoni A. ECOG performance status ≥2 as a prognostic factor in patients with advanced non small cell lung cancer treated with immune checkpoint inhibitors-A systematic review and meta-analysis of real world data. Lung Cancer. 2020;145:95–104. doi:10.1016/j.lungcan.2020.04.027

30. Rizzo MM, Bluthgen MV, Recondo G, et al. Outcomes of patients with non-small cell lung cancer and poor performance status treated with immune checkpoint inhibitors in the real-world setting. Int J Clin Oncol. 2021;26(6):1057–1064. doi:10.1007/s10147-021-01896-x

31. Sehgal K, Gill RR, Widick P, et al. Association of Performance Status With Survival in Patients With Advanced Non–Small Cell Lung Cancer Treated With Pembrolizumab Monotherapy. JAMA Netw Open. 2021;4(2):e2037120. doi:10.1001/jamanetworkopen.2020.37120

