## Supplementary for "Comprehensive real-world utilization, treatment trajectories, and outcomes of immune checkpoint inhibitors across cancer types: the multicenter FAIR-ICI cohort"

Supplementary Material

**FAIR-ICI interactive dashboard**

The interactive dashboard provides access to aggregated cohort characteristics, treatment patterns, and clinical outcomes, with dynamic subgroup filtering. Small cells are masked to protect patient privacy.

Full URL:
https://app.powerbi.com/view?r=eyJrIjoiMjQ5ZDczYjQtOThlMS00ZGM3LThlMzEtODE4ZDQxMTg0NWQ4IiwidCI6Ijk3MjYyNjI0LTZhOTctNDFjZS04NjAyLTIyZmM0M2I3MzYzYSIsImMiOjl9

#### Supplementary Methods

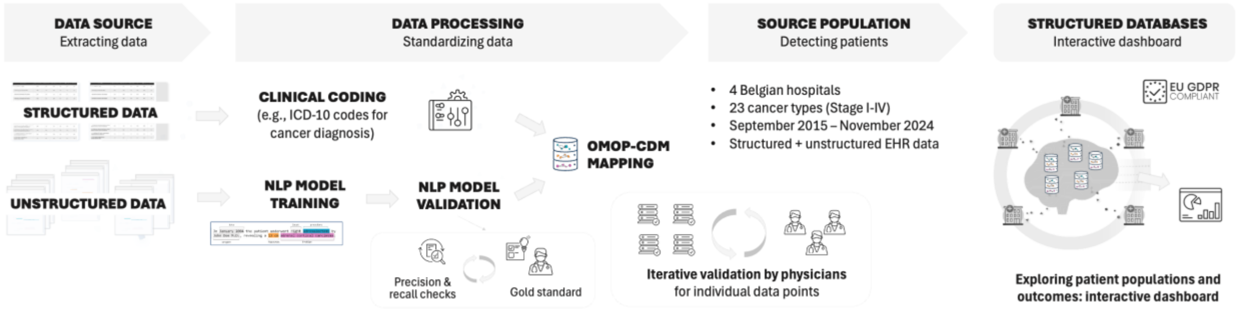

***Figure S1. Data extraction and processing workflow***

***Table S1. Data point list***

| Data point | Source | OMOP Table | Concept identifier(s) |
|---|---|---|---|
| Alcohol abuse | All | Observation | C0085762 |
| ALK TKI | Structured | Drug exposure | C5432218 |
| Antineoplastic agents | Structured | Drug exposure | C0003392 |
| Atezolizumab (Tecentriq) | Structured | Drug exposure | C4055433 |
| Avelumab (Bavencio) | Structured | Drug exposure | C4055417 |
| Axitinib | Structured | Drug exposure | C1700874, C3265883 |
| basal cell carcinoma skin | All | Condition occurrence | C4721806 |
| Bevacizumab | Structured | Drug exposure | C0796392 |
| Biliary cancer | All | Condition occurrence | C0750952 |
| Bladder cancer | All | Condition occurrence | C0005684, C0005695 |
| BMI | All | Measurement | C1305855 |
| Breast cancer | All | Condition occurrence | C0006142, C0678222 |
| Cancer | All | Condition occurrence | C1306459, C0006826 |
| Cancer stage | All | Measurement | C0804690, C1300486, C0027646, C0804696 |
| Carboplatin | Structured | Drug exposure | C0079083 |

| | | | |
|---|---|---|---|
| Cemiplimab (Libtayo) | Structured | Drug exposure | C4724806 |
| Cervix cancer | All | Condition occurrence | C4048328, C0007847 |
| Cholangiocarcinoma | All | Condition occurrence | C0206698 |
| Cisplatin | Structured | Drug exposure | C0008838 |
| Classical Hodgkin lymphoma | All | Condition occurrence | C1333064 |
| Clinical metastatic stage | All | Measurement | C1275856 |
| Clinical nodal stage | All | Measurement | C1275865 |
| Clinical tumor stage | All | Measurement | C1275863 |
| Colorectal carcinoma | All | Condition occurrence | C0009402, C4722085 |
| Death | Structured | Person | C0011065 |
| Dostarlimab (Jemperli) | Structured | Drug exposure | C5242455 |
| Durvalumab (Imfinzi) | Structured | Drug exposure | C4473954, C4055109 |
| ECOG performance status finding | All | Measurement | C1828127, C1520224 |
| EGFR TKI | Structured | Drug exposure | C5574906 |
| Endometrial cancer | All | Condition occurrence | C0014170 |
| Esophageal carcinoma | All | Condition occurrence | C0152018 |
| Etoposide | Structured | Drug exposure | C0015133 |
| Fluorouracil | Structured | Drug exposure | C0016360 |
| Gallbladder cancer | All | Condition occurrence | C0153452 |
| Gastrointestinal cancer | All | Condition occurrence | C0685938 |
| Head and neck cancer | All | Condition occurrence | C0278996 |
| Height | All | Measurement | C0005890 |
| Hepatocellular carcinoma | All | Condition occurrence | C2239176 |
| HER2 TKI | Structured | Drug exposure | C5432225 |

| | | | |
|---|---|---|---|
| Ipilimumab (Yervoy) | Structured | Drug exposure | C1367202 |
| Karnofsky Performance Status | All | Measurement | C4696566, C4545792, C0206065 |
| Lung cancer | All | Condition occurrence | C0242379 |
| Malignant mesothelioma | All | Condition occurrence | C0345967 |
| Malignant neoplasm of stomach | All | Condition occurrence | C0024623 |
| Melanoma | All | Condition occurrence | C0346040, C0025202 |
| Merkel cell carcinoma | All | Condition occurrence | C0007129 |
| Metastatic neoplasm | All | Condition occurrence | C2939420, C2939419 |
| Metastatic stage | All | Measurement | C0456533 |
| Nivolumab (Opdivo) | Structured | Drug exposure | C3657270 |
| Nodal stage | All | Measurement | C0456532 |
| Non-small cell lung cancer | All | Condition occurrence | C0345958, C0152013, C0007131, C0149782 |
| Ovarian cancer | All | Condition occurrence | C1140680 |
| Paclitaxel | Structured | Drug exposure | C0144576 |
| Pathological metastatic stage | All | Measurement | C1269798 |
| Pathological nodal stage | All | Measurement | C1269796 |
| Pathological tumor stage | All | Measurement | C0332389, C1270004 |
| Patient date of birth | Structured | Person | C0421451 |
| Patient sex | Structured | Person | C0079399 |
| Pazopanib | Structured | Drug exposure | C1831796 |
| Pembrolizumab (Keytruda) | Structured | Drug exposure | C3855203, C3658706 |
| Pemetrexed (Alimta) | Structured | Drug exposure | C0210657 |
| Prostate cancer | All | Condition occurrence | C0376358 |

| | | | |
|---|---|---|---|
| Renal cell carcinoma | All | Condition occurrence | C0279702, C0007134, C1266044 |
| Small cell lung cancer | All | Condition occurrence | C0149925 |
| Sorafenib | Structured | Drug exposure | C1516119 |
| Squamous cell carcinoma skin | All | Condition occurrence | C0553723 |
| Sunitinib | Structured | Drug exposure | C1176020 |
| TNM category | All | Measurement | C0809869 |
| Tobacco smoking status | All | Observation | C0337671, C0337672, C0337667, C0337664, C0425310, C0453996, C0425293 |
| Tremelimumab (Imjudo) | Structured | Drug exposure | AT36860767, LC5500009, C2351038 |
| Tumor stage | All | Measurement | C0475455 |
| Urothelial carcinoma | All | Condition occurrence | C4288754, C0279680, C0007138 |
| VEGFR TKI | Structured | Drug exposure | C5432233 |
| Weight | All | Measurement | C0005910 |
| WHO performance status | All | Measurement | C1298651 |

***Table S2. Data dictionary***

| **Variable** | **Datapoint (or variable)** | **OMOP table** | **Calculation logic** | **Source** |
|---|---|---|---|---|
| Index date (ICI initiation) | ICI administration | Drug exposure | Defined as the first recorded administration of an ICI. Only the first ICI initiation per patient is retained. Timing: first administration within the study observation period. | Structured |
| Cancer type (ICI indication) | Cancer diagnosis | Condition occurrence | Identified using structured MTB and MZG records and NLP-derived diagnoses. Assigned based on source agreement, timing, and reimbursed indications; ambiguous or unidentified indications classified as unknown. See Supplementary Methods S1 for | Structured + NLP |

| | | | | |
|---|---|---|---|---|
| | | | the detailed assignment algorithm. | |
| Primary diagnosis date | Cancer diagnosis | Condition occurrence | Determined from the source used to assign the ICI indication. Structured MTB or MZG diagnosis dates used where applicable; otherwise the earliest relevant NLP-derived diagnosis date or report date. | Structured + NLP |
| Cancer stage | Cancer stage | Measurement | Derived using UICC TNM 8th edition. Directly recorded stage used when available; otherwise calculated from TNM, prioritizing pathological, clinical, then general TNM. Timing: at diagnosis and index; index window [−120, +60] days. See Supplementary Methods S2 for the detailed staging algorithm. | Structured + NLP |
| Pathological TNM | Pathological tumor / nodal / metastatic stage | Measurement | Extracted from structured data or NLP; preferentially used for staging. Timing: structured staging at diagnosis; NLP closest within [−60, +60] days of diagnosis or [−120, +60] days of index. | Structured + NLP |
| Clinical TNM | Clinical tumor / nodal / metastatic stage | Measurement | Used when pathological TNM is unavailable. Timing: closest within [−60, +60] days of diagnosis or [−120, +60] days of index. | Structured + NLP |
| General TNM | Tumor / nodal / metastatic stage | Measurement | Consolidated TNM values, prioritizing pathological over clinical T and N; highest M retained. If pM = 0 and cM is unavailable, M is missing. Timing: corresponding staging windows. | Structured + NLP |
| Metastatic status at index | Cancer stage / Metastatic neoplasm | Measurement / Condition occurrence | Stage IV or documented metastatic disease at index, using structured records and NLP-derived metastasis mentions. Timing: before index or within [−120, +60] days. | Structured + NLP |
| Performance status | ECOG / WHO / Karnofsky | Measurement | Closest performance status selected; Karnofsky converted to ECOG using the Belgian Cancer Registry conversion matrix. For ambiguous scores, | Structured + NLP |

| | | | | |
|---|---|---|---|---|
| | | | first value retained. Timing: [−60, +60] days around index. | |
| Age at index | Birth date / Index date | Person | Calculated from date of birth and ICI initiation date; subsequently categorized for privacy preservation. | Structured |
| Sex | Gender concept ID | Person | Derived from the recorded OMOP gender concept ID. | Structured |
| Weight at index | Weight | Measurement | Closest valid weight selected; values outside 25–300 kg excluded. Timing: [−60, +30] days around index. | Structured + NLP |
| Height at index | Height | Measurement | Closest available height; units standardized and values outside 130–250 cm excluded. No fixed timing restriction. | Structured + NLP |
| Body mass index | BMI / Weight / Height | Measurement | Closest recorded BMI or BMI calculated from weight and height. Values outside 10–70 $kg/m^2$ excluded. Timing: [−60, +30] days around index. | Structured + NLP |
| Smoking status | Tobacco smoking status | Measurement / Condition occurrence / Observation | Closest recorded status categorized as non-smoker, former smoker, current smoker, or unknown. Timing: on or before index. | Structured + NLP |
| Alcohol abuse | Alcohol abuse | Measurement / Condition occurrence / Observation | Documented alcohol abuse; absence of documentation classified as unknown. Timing: on or before index. | Structured + NLP |
| Systemic treatment | Antineoplastic agents | Drug exposure | Antineoplastic drugs identified using the ATHENA concept hierarchy. Timing: recorded administrations during observation. | Structured |
| Chemotherapy | Cytotoxic antineoplastic agents | Drug exposure | Identified using antineoplastic drug concepts; administrations grouped into drug eras allowing gaps of ≤30 days. | Structured |
| Immunotherapy (ICI) | Nivolumab / Pembrolizumab / Cemiplimab / Dostarlimab / Atezolizumab / Durvalumab / Avelumab / Ipilimumab / Tremelimumab | Drug exposure | First administration defines index. Subsequent administrations grouped into drug eras. Timing: from index throughout follow-up. | Structured |
| Targeted therapy | Tyrosine kinase inhibitors (TKIs) | Drug exposure | Identified using predefined TKI drug concepts, including | Structured |

| | | | | |
|---|---|---|---|---|
| | | | agents used in combination with ICIs. | |
| VEGF inhibitors | Bevacizumab / Ramucirumab | Drug exposure | Identified from recorded drug administrations during treatment observation. | Structured |
| Drug era | Systemic treatment | Drug exposure | Continuous exposure periods defined by grouping administrations separated by ≤30 days. | Structured |
| Regimen | Drug era | Derived | Constructed by combining overlapping or closely timed drug eras (≤30-day separation). | Structured |
| Line of treatment | Regimen | Derived | New treatment line assigned when a regimen changes and at least one new drug is introduced. | Structured |
| Permanent ICI discontinuation | ICI administration | Drug exposure / Derived | Identified by a gap of at least 12 weeks without ICI reinitiation. Last ICI administration date retained. | Structured |
| Overall survival (OS) | Index date / Death date | Derived | Time from ICI initiation to death; censored at last known follow-up or applicable cohort exit. | Structured |
| Time on treatment (ToT) | ICI administration / Regimen | Derived | Time from ICI initiation to permanent discontinuation, initiation of a new treatment regimen, or death. | Structured |
| Time to next treatment (TTNT) | Regimen | Derived | Time from ICI initiation to the start of the next treatment regimen; censored at end of observation if no subsequent regimen is recorded. | Structured |

***Table S3. NLP metrics of relevant variables***

| Variable | Precision | Precision n | Recall | Recall n | F1 Score |
|---|---|---|---|---|---|
| Adjuvant therapy | 100.0% | 27 | 93.8% | 16 | 96.8% |
| Alcohol abuse | 100.0% | 5 | 93.8% | 16 | 96.8% |
| BMI | NA | NA | 100.0% | 8 | NA |
| Basal cell carcinoma skin | NA | NA | 100.0% | 4 | NA |
| Breast cancer | 97.1% | 240 | 90.2% | 50 | 93.5% |
| Cancer | 97.3% | 629 | 92.7% | 620 | 94.9% |
| Cancer progression | 99.3% | 138 | 96.2% | 26 | 97.7% |
| Cancer relapse | 100.0% | 63 | 66.7% | 3 | 80.0% |
| Cancer stage | 97.6% | 127 | 88.1% | 42 | 92.6% |

| | | | | | |
|---|---|---|---|---|---|
| Cervix cancer | 100.0% | 2 | NA | NA | NA |
| Classical Hodgkin lymphoma | 100.0% | 50 | NA | NA | NA |
| Clinical metastatic stage | 100.0% | 26 | 94.5% | 73 | 97.2% |
| Clinical nodal stage | 100.0% | 16 | 89.8% | 88 | 94.6% |
| Clinical tumor stage | 98.0% | 101 | 95.3% | 85 | 96.6% |
| Colorectal carcinoma | 98.1% | 52 | 100.0% | 2 | 99.0% |
| ECOG performance status finding | 94.0% | 50 | 100.0% | 10 | 96.9% |
| Endometrial cancer | 100.0% | 50 | 100.0% | 1 | 100.0% |
| Esophageal carcinoma | 100.0% | 50 | 100.0% | 1 | 100.0% |
| Fatigue | 95.0% | 20 | 97.2% | 71 | 96.1% |
| Gastrointestinal cancer | 94.1% | 17 | 100.0% | 1 | 97.0% |
| Head and neck cancer | 96.6% | 58 | 100.0% | 3 | 98.3% |
| Height | 100.0% | 4 | 100.0% | 10 | 100.0% |
| Hepatocellular carcinoma | NA | NA | 100.0% | 1 | NA |
| Lung cancer | 96.9% | 192 | 100.0% | 90 | 98.4% |
| Malignant mesothelioma | NA | NA | 80.0% | 5 | NA |
| Melanoma | 98.8% | 164 | 100.0% | 8 | 99.4% |
| Merkel cell carcinoma | 100.0% | 50 | NA | NA | NA |
| Metastatic neoplasm | 97.2% | 288 | 94.8% | 193 | 96.0% |
| Metastatic stage | 97.8% | 91 | 89.6% | 48 | 93.5% |
| Neoadjuvant therapy | 98.2% | 57 | NA | NA | NA |
| Nodal stage | 97.3% | 187 | 82.9% | 70 | 89.5% |
| Non-small cell lung cancer | 98.8% | 167 | 94.0% | 67 | 96.3% |
| Ovarian cancer | NA | NA | 100.0% | 2 | NA |
| Pathological M-stage neo-adj | 100.0% | 2 | 100.0% | 1 | 100.0% |
| Pathological metastatic stage | 100.0% | 9 | 94.3% | 53 | 97.1% |
| Pathological nodal stage | 100.0% | 20 | 92.9% | 85 | 96.3% |
| Pathological nodal stage neo-adj | 100.0% | 3 | 42.9% | 7 | 60.0% |
| Pathological tumor stage | 100.0% | 37 | 94.8% | 97 | 97.3% |
| Pathological tumor stage neo-adj | 100.0% | 4 | 70.0% | 10 | 82.4% |
| Progressive disease | 100.0% | 9 | 96.9% | 32 | 98.4% |
| Prostate cancer | 100.0% | 51 | 100.0% | 2 | 100.0% |
| Radiotherapy | 98.3% | 174 | 95.5% | 156 | 96.9% |
| Renal cell carcinoma | 100.0% | 50 | 100.0% | 5 | 100.0% |
| Resection | 100.0% | 33 | 84.2% | 38 | 91.4% |
| Small-cell lung cancer | 100.0% | 53 | 100.0% | 21 | 100.0% |
| Stable disease | 100.0% | 56 | 90.0% | 10 | 94.7% |

| | | | | | |
|---|---|---|---|---|---|
| TNM category | 85.7% | 7 | 100.0% | 9 | 92.3% |
| Tobacco smoking status | 86.4% | 66 | 100.0% | 65 | 92.7% |
| Tumor grade | 100.0% | 13 | 87.5% | 24 | 93.3% |
| Tumor stage | 100.0% | 64 | 89.1% | 64 | 94.2% |
| Urothelial carcinoma | 95.0% | 100 | 100.0% | 9 | 97.4% |
| Weight | 100.0% | 10 | 95.8% | 24 | 97.9% |
| WHO performance status | 100.0% | 96 | 100.0% | 33 | 100.0% |

Metrics are provided by Lynxcare Health Informatics. Clinical validation was done independently by physicians participating in the FAIR-ICI study (AV, CV, ADM, PRD, SA, MV).

***Methods S1. Cancer Type and ICI Indication Assignment Algorithm***

Cancer type was derived from three sources: structured hospital cancer registry (ICD-O-3), minimal hospital data (ICD-10), and unstructured clinical text processed through NLP. For each source, we evaluated whether the administered ICI was reimbursed for the corresponding tumor type. When multiple sources were available, cancer type and reimbursed indication were assigned based on concordance across sources; when discordant, priority was given to cancer registry, followed by minimal hospital data and NLP.

Indication assignment followed these rules:

1. Cancer registry–NLP concordance: The same tumor type was identified in cancer registry and NLP; this match was used for indication assignment.
2. Minimal hospital data–NLP concordance: The same tumor type was identified in minimal hospital data and NLP; this match was used for indication assignment.
3. No concordance across sources: When structured and unstructured data did not align, tumor type was assigned based on an individual source according to the predefined hierarchy.
4. More than one reimbursed indication: Disambiguation was performed using treatment regimen information or manual review.

Belgian reimbursement indications were used to support cancer-type assignment. Patients without an identifiable matching reimbursed indication, potentially including those treated through clinical trials or compassionate use, or with multiple unresolved possible indications were classified as having an unknown cancer type.

The full logic is illustrated in Figure S2, with patient counts by indication assignment source presented in Table S4.

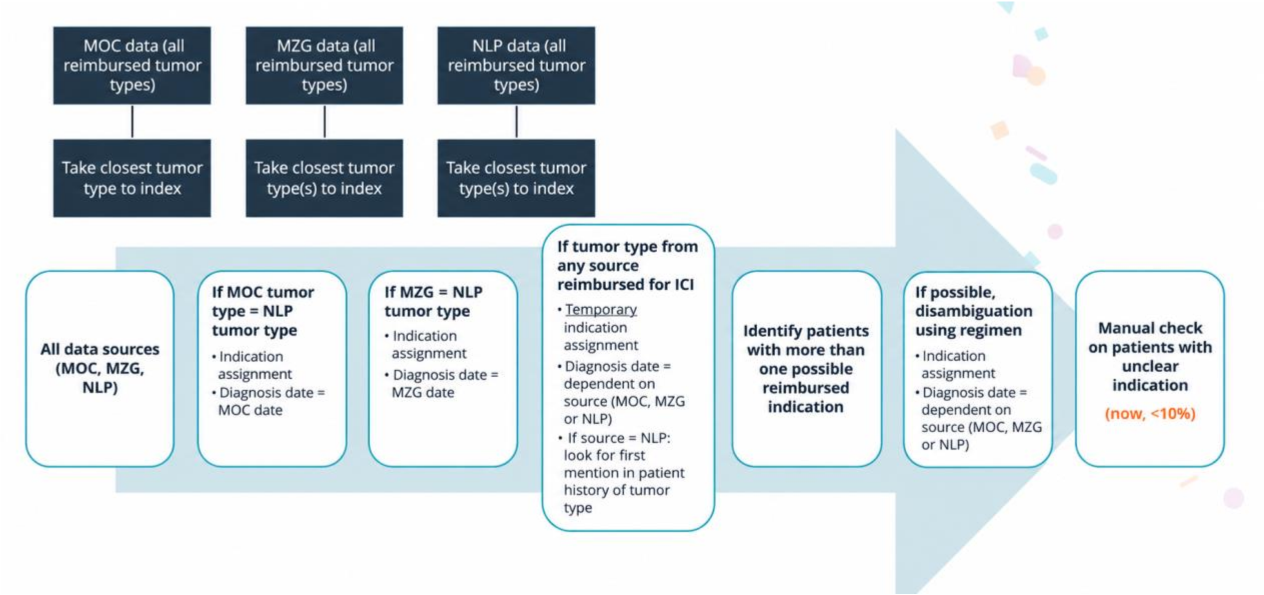

***Figure S2. Cancer type and indication assignment Logic***. MOC = local hospital cancer registry. MZG = minimal hospital data.

***Table S4. Source used for cancer type and ICI indication assignment***

| **Source/assignment scenario** | **Patients, n (%)** |
|---|---|
| Cancer registry | 373 (11.5%) |
| Cancer registry–NLP concordance | 1,883 (58.3%) |
| Minimal hospital data | 74 (2.3%) |
| Minimal hospital data–NLP concordance | 400 (12.4%) |
| NLP only | 215 (6.7%) |
| Indication unclear | 286 (8.9%) |
| **Total** | **3,231 (100%)** |

***Methods S2. Tumor Staging Algorithm***

Tumor stage at diagnosis and at the index date was determined according to the Union for International Cancer Control (UICC) TNM Classification (8th edition). When multiple TNM records were available for a patient, pathological TNM (pTNM) was given priority, followed by clinical TNM (cTNM), and then unspecified TNM entries.

For staging at diagnosis, patients were classified as Stage IV whenever a metastasis-related record was identified during the period extending from 60 days before to 60 days after the diagnosis date. The terms considered indicative of metastasis included “metastatic neoplasm”, “cancer relapse”, and “cancer progression”, while terms referring solely to lymph-node involvement or local recurrence (such as “klier”, “lokaal recidief”, or ICD code C77) were explicitly excluded. When this criterion was met, the Stage IV assignment overrode any available TNM information.

Among patients who did not meet the metastasis criterion, stage at diagnosis was assigned using a hierarchical approach. First, stage explicitly recorded in the cancer registry was used. If not available, stage was calculated from TNM values recorded in the cancer registry. If neither was present, stage extracted directly from NLP-processed clinical text was applied. Finally, if needed, stage was calculated from TNM values derived through NLP. When staging was based on TNM, missing T, N, or M components were tolerated provided that the missing information did not alter the final stage classification; for example, in breast cancer, any T with N3 and M0 was treated as Stage III regardless of the specific T value.

Stage at ICI initiation was determined using staging information within 120 days before to 60 days after ICI initiation, applying the TNM hierarchy and incorporating metastasis-related records. Patients were considered metastatic at ICI initiation if Stage IV disease was identified or a metastatic concept was documented before index or at cancer diagnosis.

### Supplementary Results

***Table S5. Baseline characteristics at ICI initiation, in other known cancer types***

| | **Lung cancer unspecified** | **Breast cancer** | **Small cell lung cancer (SCLC)** | **Meso-thelioma** | **Hepato-cellular carcinoma** | **Biliary cancer** | **Cervical cancer** | **Colorectal cancer** |
|---|---|---|---|---|---|---|---|---|
| | (N=184) | (N=146) | (N=135) | (N=80) | (N=76) | (N=41) | (N=38) | (N=37) |
| **Demographics** | | | | | | | | |
| **Age** | | | | | | | | |
| 18-49 | 8 (4.3%) | 53 (36.3%) | 6 (4.4%) | <5 | 7 (9.2%) | <5 | 11 (28.9%) | <5 |
| 50-64 | 65 (35.3%) | 50 (34.2%) | 44 (32.6%) | 18 (22.5%) | 13 (17.1%) | 16 (39.0%) | 20 (52.6%) | 8 (21.6%) |
| 65-74 | 70 (38.0%) | 27 (18.5%) | 63 (46.7%) | 44 (55.0%) | 28 (36.8%) | 20 (48.8%) | 5 (13.2%) | 11 (29.7%) |
| ≥75 | 41 (22.3%) | 16 (11.0%) | 22 (16.3%) | 14 (17.5%) | 28 (36.8%) | <5 | <5 | 14 (37.8%) |
| **Sex** | | | | | | | | |
| Female | 53 (28.8%) | 145 (99.3%) | 47 (34.8%) | 18 (22.5%) | 13 (17.1%) | 15 (36.6%) | 38 (100%) | 24 (64.9%) |
| Male | 131 (71.2%) | <5 | 88 (65.2%) | 62 (77.5%) | 63 (82.9%) | 26 (63.4%) | - | 13 (35.1%) |
| **BMI category** | | | | | | | | |
| Normal | 66 (35.9%) | 56 (38.4%) | 37 (27.4%) | 25 (31.3%) | 14 (18.4%) | 9 (22.0%) | 17 (44.7%) | 14 (37.8%) |
| Underweight | 18 (9.8%) | 8 (5.5%) | 7 (5.2%) | <5 | <5 | <5 | <5 | <5 |
| Overweight | 37 (20.1%) | 34 (23.3%) | 47 (34.8%) | 33 (41.3%) | 25 (32.9%) | 12 (29.3%) | 6 (15.8%) | 8 (21.6%) |
| Obese | 22 (12.0%) | 33 (22.6%) | 26 (19.3%) | 11 (13.8%) | 23 (30.3%) | 7 (17.1%) | 6 (15.8%) | 5 (13.5%) |
| Unknown | 41 (22.3%) | 15 (10.3%) | 18 (13.3%) | 8 (10.0%) | 12 (15.8%) | 11 (26.8%) | 8 (21.1%) | 9 (24.3%) |
| **Clinical / Lifestyle Characteristics** | | | | | | | | |
| **Performance status (ECOG)** | | | | | | | | |
| 0-1 | 28 (15.2%) | 45 (30.8%) | 41 (30.4%) | 33 (41.3%) | 31 (40.8%) | 12 (29.3%) | 19 (50.0%) | <5 |
| ≥2 | 23 (12.5%) | 5 (3.4%) | 16 (11.9%) | 6 (7.5%) | 9 (11.8%) | 5 (12.2%) | <5 | <5 |
| Unknown | 133 (72.3%) | 96 (65.8%) | 78 (57.8%) | 41 (51.3%) | 36 (47.4%) | 24 (58.5%) | 17 (44.7%) | 31 (83.8%) |
| **Smoking status** | | | | | | | | |
| Non-smoker | 13 (7.1%) | 34 (23.3%) | 7 (5.2%) | 26 (32.5%) | 24 (31.6%) | 10 (24.4%) | 13 (34.2%) | 5 (13.5%) |
| Smoker | 92 (50.0%) | 19 (13.0%) | 67 (49.6%) | 17 (21.3%) | 17 (22.4%) | 8 (19.5%) | 9 (23.7%) | 5 (13.5%) |
| Ex-smoker | 21 (11.4%) | 12 (8.2%) | 33 (24.4%) | 15 (18.8%) | 9 (11.8%) | <5 | <5 | <5 |
| Unknown | 58 (31.5%) | 81 (55.5%) | 28 (20.7%) | 22 (27.5%) | 26 (34.2%) | 19 (46.3%) | 13 (34.2%) | 24 (64.9%) |
| **Alcohol abuse** | | | | | | | | |
| No/Unknown | 157 (85.3%) | 143 (97.9%) | 114 (84.4%) | 78 (97.5%) | 45 (59.2%) | 34 (82.9%) | 37 (97.4%) | 32 (86.5%) |
| Yes | 27 (14.7%) | <5 | 21 (15.6%) | <5 | 31 (40.8%) | 7 (17.1%) | <5 | 5 (13.5%) |
| **Disease characteristics** | | | | | | | | |
| **Metastatic status at ICI start** | | | | | | | | |
| Non-metastatic | 8 (4.3%) | 66 (45.2%) | - | - | - | <5 | - | <5 |
| Metastatic | 161 (87.5%) | 67 (45.9%) | 133 (98.5%) | 79 (98.8%) | 72 (94.7%) | 36 (87.8%) | 36 (94.7%) | 30 (81.1%) |
| Unknown | 15 (8.2%) | 13 (8.9%) | <5 | <5 | <5 | <5 | <5 | <5 |
| **Stage at ICI start** | | | | | | | | |
| Stage I | <5 | 10 (6.8%) | <5 | 6 (7.5%) | - | <5 | - | - |
| Stage II | <5 | 43 (29.5%) | - | - | 5 (6.6%) | <5 | - | <5 |
| Stage III | 6 (3.3%) | 15 (10.3%) | <5 | 15 (18.8%) | 6 (7.9%) | <5 | <5 | <5 |
| Stage IV | 160 (87.0%) | 66 (45.2%) | 130 (96.3%) | 45 (56.3%) | 53 (69.7%) | 35 (85.4%) | 36 (94.7%) | 29 (78.4%) |
| Unknown | 15 (8.2%) | 12 (8.2%) | <5 | 14 (17.5%) | 12 (15.8%) | <5 | <5 | <5 |
| **Treatment patterns** | | | | | | | | |
| **ICI mechanisms** | | | | | | | | |
| Anti-PD-1 | 129 (70.1%) | 125 (85.6%) | 9 (6.7%) | 30 (37.5%) | - | <5 | 38 (100%) | 37 (100%) |
| Anti-PD-L1 | 50 (27.2%) | 21 (14.4%) | 126 (93.3%) | - | 76 (100%) | 38 (92.7%) | - | - |
| Anti-CTLA-4 | - | - | - | - | - | - | - | - |
| Anti-PD-1 + Anti-CTLA-4 | 5 (2.7%) | - | - | 50 (62.5%) | - | - | - | - |
| **Treatment regimen group** | | | | | | | | |
| ICI mono | 104 (56.5%) | 20 (13.7%) | 15 (11.1%) | 30 (37.5%) | 19 (25.0%) | <5 | 18 (47.4%) | 34 (91.9%) |
| ICI dual | 5 (2.7%) | - | - | 50 (62.5%) | - | - | - | - |
| ICI + chemo | 73 (39.7%) | 125 (85.6%) | 119 (88.1%) | - | <5 | 37 (90.2%) | 19 (50.0%) | <5 |
| ICI + TKI | <5 | <5 | <5 | - | <5 | - | - | <5 |
| ICI + other | <5 | - | - | - | 54 (71.1%) | - | <5 | - |

Table S5 (continued)

| | gastric cancer | endometrial cancer | cutaneous squamous cell carcinoma | esophageal adeno-carcinoma | merckel cell carcinoma | basal cell carcinoma | esophageal squamous cell carcinoma | hodgkin lymphoma |
|---|---|---|---|---|---|---|---|---|
| | (N=36) | (N=34) | (N=30) | (N=19) | (N=14) | (N=8) | (N=8) | (N=7) |
| **Demographics** | | | | | | | | |
| **Age** | | | | | | | | |
| 18-49 | <5 | <5 | - | <5 | - | - | <5 | 6 (85.7%) |
| 50-64 | 13 (36.1%) | 10 (29.4%) | <5 | 5 (26.3%) | <5 | <5 | - | - |
| 65-74 | 11 (30.6%) | 10 (29.4%) | 8 (26.7%) | 7 (36.8%) | <5 | <5 | <5 | - |
| ≥75 | 8 (22.2%) | 13 (38.2%) | 19 (63.3%) | 5 (26.3%) | 8 (57.1%) | 5 (62.5%) | <5 | <5 |
| **Sex** | | | | | | | | |
| Female | 14 (38.9%) | 34 (100%) | 13 (43.3%) | <5 | 5 (35.7%) | <5 | <5 | <5 |
| Male | 22 (61.1%) | - | 17 (56.7%) | 16 (84.2%) | 9 (64.3%) | <5 | <5 | <5 |
| **BMI category** | | | | | | | | |
| Normal | 20 (55.6%) | 6 (17.6%) | 7 (23.3%) | 6 (31.6%) | 6 (42.9%) | <5 | <5 | <5 |
| Underweight | <5 | - | <5 | <5 | - | - | - | <5 |
| Overweight | 6 (16.7%) | 13 (38.2%) | 6 (20.0%) | 6 (31.6%) | <5 | <5 | - | <5 |
| Obese | <5 | 10 (29.4%) | 6 (20.0%) | <5 | 6 (42.9%) | <5 | - | <5 |
| Unknown | 8 (22.2%) | 5 (14.7%) | 9 (30.0%) | <5 | <5 | <5 | 7 (87.5%) | - |
| **Clinical / Lifestyle Characteristics** | | | | | | | | |
| **Performance status (ECOG)** | | | | | | | | |
| 0-1 | 9 (25.0%) | 14 (41.2%) | 6 (20.0%) | 6 (31.6%) | <5 | <5 | - | - |
| ≥2 | <5 | 8 (23.5%) | <5 | <5 | - | - | - | - |
| Unknown | 23 (63.9%) | 12 (35.3%) | 22 (73.3%) | 12 (63.2%) | 10 (71.4%) | 7 (87.5%) | 8 (100%) | 7 (100%) |
| **Smoking status** | | | | | | | | |
| Non-smoker | 5 (13.9%) | 17 (50.0%) | <5 | <5 | <5 | <5 | - | <5 |
| Smoker | <5 | <5 | <5 | 8 (42.1%) | <5 | <5 | - | <5 |
| Ex-smoker | <5 | <5 | 5 (16.7%) | <5 | <5 | <5 | <5 | <5 |
| Unknown | 25 (69.4%) | 12 (35.3%) | 19 (63.3%) | 5 (26.3%) | 9 (64.3%) | <5 | 7 (87.5%) | <5 |
| **Alcohol abuse** | | | | | | | | |
| No/Unknown | 35 (97.2%) | 34 (100%) | 30 (100%) | 15 (78.9%) | 14 (100%) | 8 (100%) | 7 (87.5%) | 7 (100%) |
| Yes | <5 | - | - | <5 | - | - | <5 | - |
| **Disease characteristics** | | | | | | | | |
| **Metastatic status at ICI start** | | | | | | | | |
| Non-metastatic | - | - | 5 (16.7%) | - | <5 | - | <5 | - |
| Metastatic | 34 (94.4%) | 32 (94.1%) | 16 (53.3%) | 15 (78.9%) | 10 (71.4%) | <5 | <5 | <5 |
| Unknown | <5 | <5 | 9 (30.0%) | <5 | <5 | <5 | <5 | <5 |
| **Stage at ICI start** | | | | | | | | |
| Stage I | - | - | - | - | <5 | - | - | - |
| Stage II | - | <5 | <5 | <5 | <5 | - | <5 | - |
| Stage III | - | - | <5 | <5 | <5 | - | - | - |
| Stage IV | 34 (94.4%) | 32 (94.1%) | 20 (66.7%) | 16 (84.2%) | 9 (64.3%) | <5 | 5 (62.5%) | <5 |
| Unknown | <5 | <5 | 5 (16.7%) | - | <5 | <5 | <5 | <5 |
| **Treatment patterns** | | | | | | | | |
| **ICI mechanisms** | | | | | | | | |
| Anti-PD-1 | 36 (100%) | 34 (100%) | 30 (100%) | 19 (100%) | - | 8 (100%) | 8 (100%) | 7 (100%) |
| Anti-PD-L1 | - | - | - | - | 14 (100%) | - | - | - |
| Anti-CTLA-4 | - | - | - | - | - | - | - | - |
| Anti-PD-1 + Anti-CTLA-4 | - | - | - | - | - | - | - | - |
| **Treatment regimen group** | | | | | | | | |
| ICI mono | 19 (52.8%) | 11 (32.4%) | 28 (93.3%) | 11 (57.9%) | 13 (92.9%) | 8 (100%) | 6 (75.0%) | 6 (85.7%) |
| ICI dual | - | - | - | - | - | - | - | - |
| ICI + chemo | 16 (44.4%) | <5 | <5 | 8 (42.1%) | <5 | - | <5 | <5 |
| ICI + TKI | <5 | 19 (55.9%) | <5 | - | - | - | - | - |
| ICI + other | - | - | - | - | - | - | - | - |
| Gallbladder carcinoma (n=1) was not presented. | | | | | | | | |

***Table S6. Outcome maturity and ICI treatment exposure by cancer type in metastatic patients receiving ICIs***

| **Cancer type** | **n** | **OS events, n (%)** | **Median follow-up, months (IQR)** | **Median follow-up among alive, months (IQR)** | **Median number of ICI administrations (IQR)** |
|---|---|---|---|---|---|
| NSCLC | 897 | 519 (57.9) | 10.7 (4–24.5) | 23.5 (9.6–43.4) | 6 (3–15) |
| Melanoma | 297 | 97 (32.7) | 20.8 (6.1–40.5) | 31.5 (19.4–56.1) | 5 (2–13) |
| Head and neck cancer | 215 | 142 (66) | 7.6 (3.1–17.5) | 14.2 (5.8–29.1) | 5 (3–12) |
| Urothelial carcinoma | 182 | 116 (63.7) | 7.8 (2.8–17.8) | 14 (5.9–26.8) | 5 (2–10) |
| Renal cell carcinoma | 190 | 83 (43.7) | 17.6 (5.7–30.1) | 21.5 (10.2–33.8) | 5 (3–17) |
| Lung cancer | 161 | 103 (64) | 6.1 (2.4–15.6) | 8.5 (2.6–32.1) | 5 (2–10) |
| Breast cancer | 67 | 22 (32.8) | 15.2 (8.4–21) | 15.8 (7.5–20.7) | 8 (5–15) |
| SCLC | 133 | 90 (67.7) | 7.2 (2.9–12.6) | 7.4 (2.9–18.5) | 6 (3–13) |
| Mesothelioma | 79 | 50 (63.3) | 9.4 (4.6–20.5) | 15.2 (5.8–25.2) | 5 (2.5–9) |
| Hepatocellular carcinoma | 72 | 37 (51.4) | 6.9 (1.9–16) | 7.3 (2.6–19.9) | 4.5 (2–13.2) |
| Biliary cancer | 36 | 13 (36.1) | 6.7 (2.5–11.5) | 8.7 (4.4–16.9) | 6 (2–9.2) |
| Cervical cancer | 36 | 12 (33.3) | 7.3 (4.4–14.2) | 9.1 (4.8–15.2) | 6 (3–10) |
| Colorectal cancer | 30 | 7 (23.3) | 10.5 (5.1–15.8) | 14 (6.6–26) | 7 (3.2–13.5) |
| Gastric cancer | 34 | 13 (38.2) | 5.1 (2.4–12.8) | 5 (2.4–11.9) | 6 (3–11) |
| Endometrial cancer | 32 | 15 (46.9) | 8.9 (2.6–14.6) | 9.6 (1.4–16.1) | 5.5 (2–12.5) |
| Cutaneous squamous cell carcinoma | 16 | 6 (37.5) | 6.1 (3.7–22.2) | 15.1 (3.3–32.4) | 4 (2–12.2) |
| Esophageal adenocarcinoma | 15 | 5 (33.3) | 7.7 (5–14.1) | 12.7 (2.6–13.3) | 8 (6–14) |
| Merckel cell carcinoma | 10 | 5 (50) | 14.9 (13.2–45.4) | 53 (22.6–63) | 17 (5–107) |
| Cancer types with n<5 were not presented. | | | | | |

***Table S7. Real-world outcomes for other cancer types in metastatic patients receiving ICIs***

| **Cancer type** | **n** | **Outcome** | **Median (months) (95%CI)** | **12-month (%)** | **24-month (%)** |
|---|---|---|---|---|---|
| Lung cancer (unspecified) | 161 | OS | 11 (8–14) | 44.1 | 25.3 |
| | | TTNT | 12 (8–32) | 47.8 | 40.5 |
| | | ToT | 6 (5–7) | 24.1 | 17.0 |
| Breast cancer | 67 | OS | 28 (22–NA) | 77.7 | 63.2 |
| | | TTNT | 5 (3–12) | 35.0 | 25.7 |
| | | ToT | 3 (3–6) | 22.8 | 12.4 |
| | 133 | OS | 11 (8–12) | 36.2 | 15.0 |

| | | | | | |
|---|---|---|---|---|---|
| Small cell lung cancer (SCLC) | | TTNT | 6 (5–7) | 20.3 | 13.0 |
| | | ToT | 5 (5–6) | 11.8 | 4.9 |
| Mesothelioma | 79 | OS | 15 (10–20) | 54.5 | 27.3 |
| | | TTNT | 2 (1–5) | 30.2 | 13.2 |
| | | ToT | 2 (1–4) | 15.8 | NA |
| Hepatocellular carcinoma | 72 | OS | 15 (11–22) | 56.7 | 27.9 |
| | | TTNT | 12 (10–17) | 49.1 | 22.5 |
| | | ToT | 7 (4–11) | 25.1 | 4.8 |
| Biliary tract cancer | 36 | OS | NR | 54.5 | NA |
| | | TTNT | 9 (6–NA) | 45.1 | NA |
| | | ToT | 6 (4–NA) | 9.6 | NA |
| Cervical cancer | 36 | OS | 19 (13–NA) | 66.5 | 47.2 |
| | | TTNT | 6 (5–NA) | 16.6 | 16.6 |
| | | ToT | 6 (5–7) | 7.8 | 7.8 |
| Colorectal cancer | 30 | OS | NR | 77.6 | 70.5 |
| | | TTNT | NR | 67.8 | 67.8 |
| | | ToT | 15 (6–NA) | 52.3 | 43.6 |
| Gastric cancer | 34 | OS | 13 (9–NA) | 52.0 | 0.0 |
| | | TTNT | 7 (4–NA) | 14.5 | NA |
| | | ToT | 5 (4–10) | 12.5 | 12.5 |
| Endometrial cancer | 32 | OS | 17 (10–NA) | 51.9 | 37.1 |
| | | TTNT | 7 (3–NA) | 30.4 | 30.4 |
| | | ToT | 4 (3–8) | 20.0 | 0.0 |
| Cutaneous squamous cell carcinoma | 16 | OS | 44 (9–NA) | 55.2 | 55.2 |
| | | TTNT | NR | 75.0 | 75.0 |
| | | ToT | 11 (4–NA) | 41.5 | 20.7 |
| Esophageal adenocarcinoma | 15 | OS | 16 (8–NA) | 64.8 | 32.4 |
| | | TTNT | 11 (8–NA) | 48.1 | 48.1 |
| | | ToT | 6 (5–NA) | 17.3 | NA |
| Merkel cell carcinoma | 10 | OS | 16 (13–NA) | 78.8 | 45.0 |
| | | TTNT | NR | 66.7 | 66.7 |
| | | ToT | NR | 53.3 | 53.3 |
| Cancer types with n<5 were not presented. | | | | | |

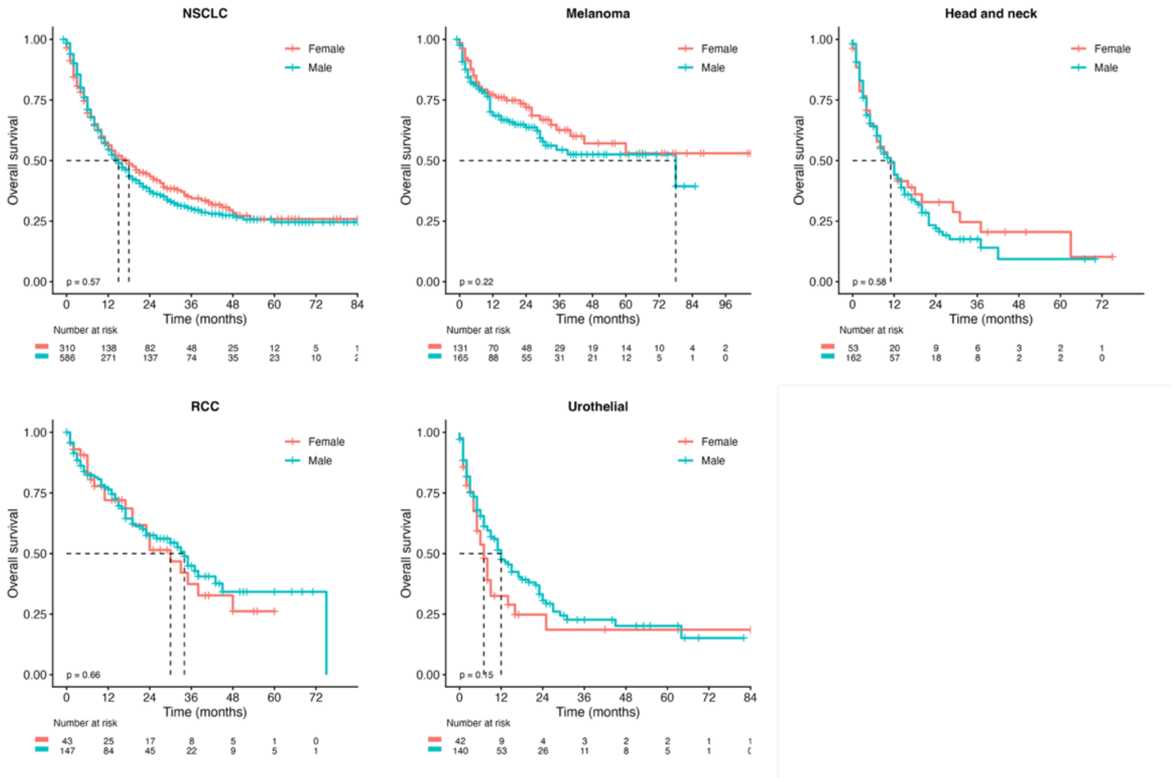

***Figure S3. Real-world overall survival stratified by sex among metastatic patients receiving ICIs, across five major cancer types***

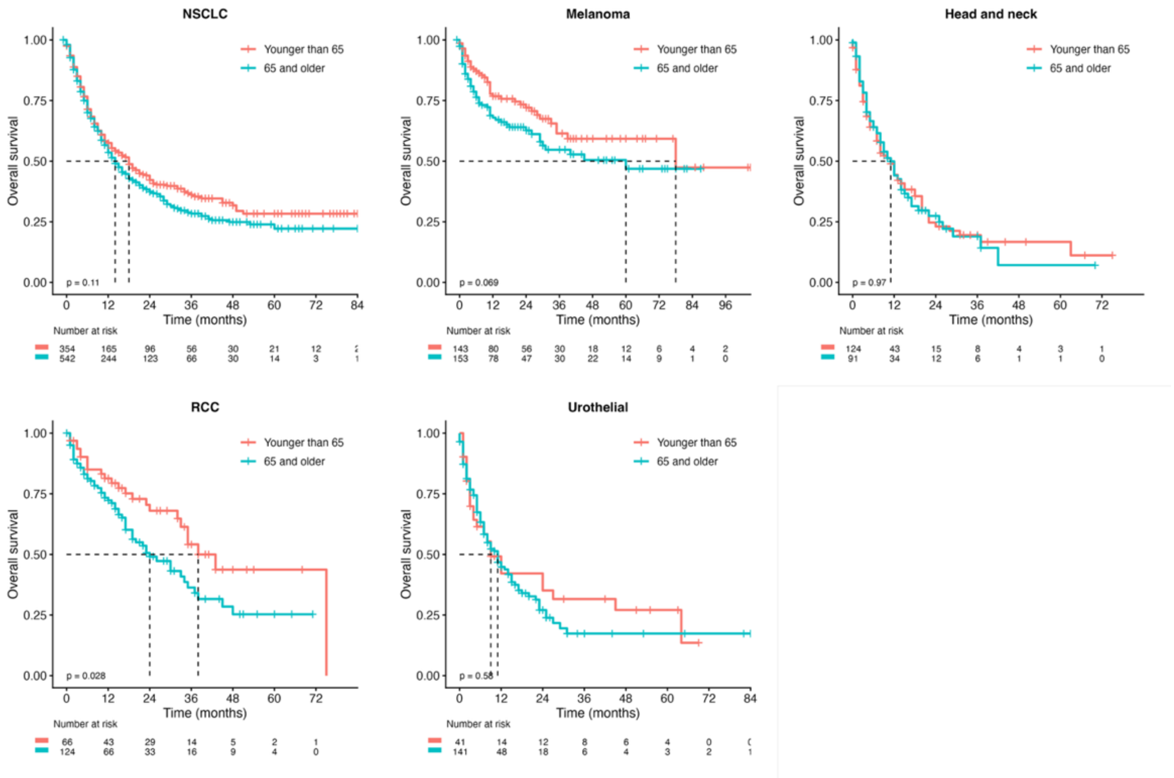

***Figure S4. Real-world overall survival stratified by age group among metastatic patients receiving ICIs, across five major cancer types***

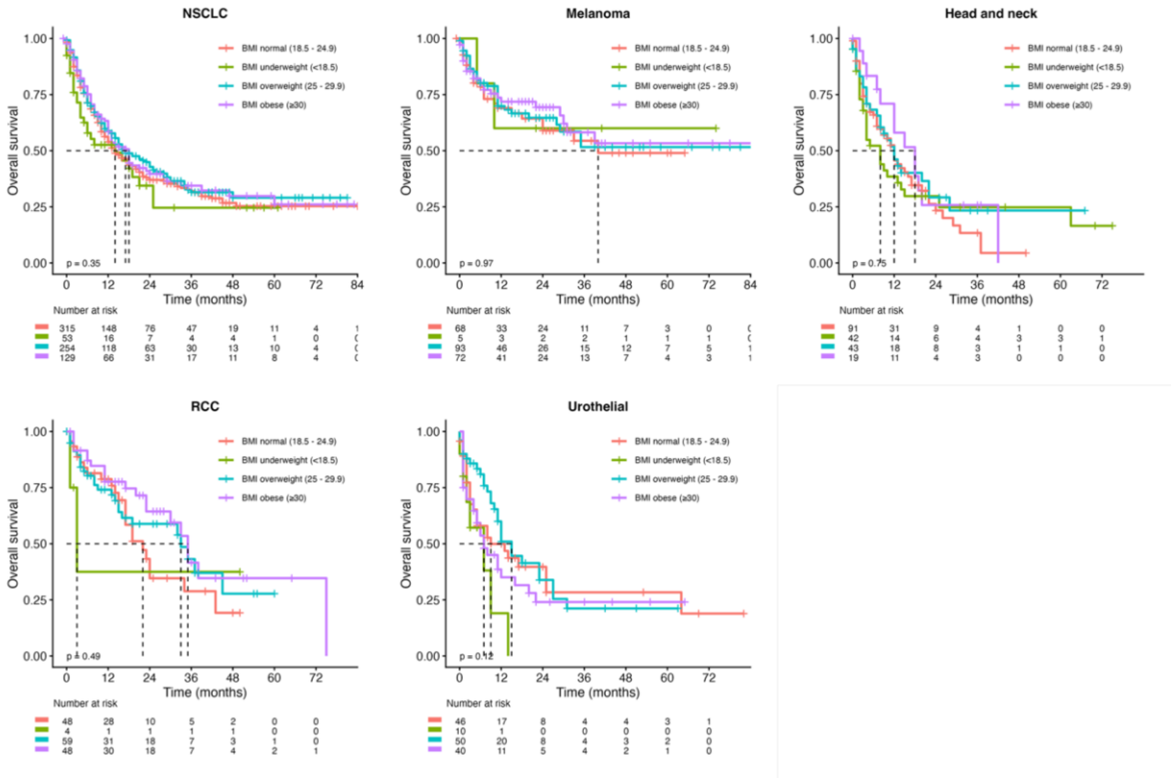

***Figure S5. Real-world overall survival stratified by BMI category among metastatic patients receiving ICIs, across five major cancer types***

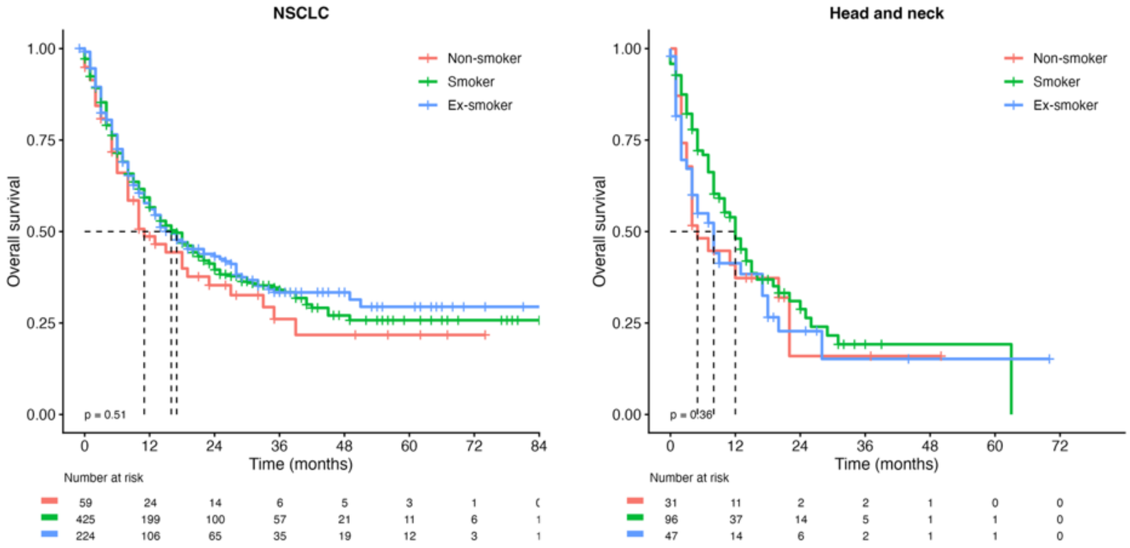

***Figure S6. Real-world overall survival stratified by smoking status among metastatic patients receiving ICIs, in NSCLC and Head and neck cancer***

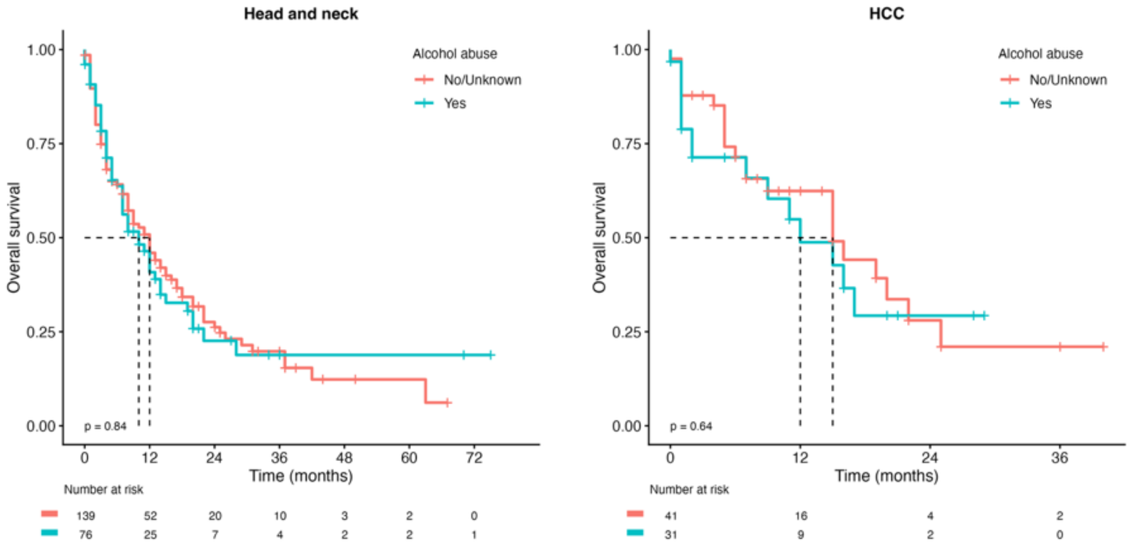

***Figure S7. Real-world overall survival stratified by alcohol use among metastatic patients receiving ICIs, in Head and neck cancer and hepatocellular carcinoma (HCC)***
